# Refined selection of individuals for preventive cardiovascular disease treatment with a Transformer-based risk model

**DOI:** 10.1101/2024.09.25.24314371

**Authors:** Shishir Rao, Yikuan Li, Mohammad Mamouei, Gholamreza Salimi-Khorshidi, Malgorzata Wamil, Milad Nazarzadeh, Christopher Yau, Gary S Collins, Rod Jackson, Andrew Vickers, Goodarz Danaei, Kazem Rahimi

**Affiliations:** Deep Medicine, Oxford Martin School, University of Oxford, Oxford, United Kingdom; Nuffield Department of Women’s & Reproductive Health, University of Oxford, Oxford, United Kingdom; Health Data Research UK, London, UK; Centre for Statistics in Medicine, Nuffield Department of Orthopaedics, Rheumatology and Musculoskeletal Sciences, Botnar Research Centre, University of Oxford, Oxford, United Kingdom; School of Population Health, Faculty of Medical and Health Sciences, University of Auckland, Auckland, New Zealand; Memorial Sloan Kettering Cancer Center, New York, United States of America; Departments of Global Health and Population, and Epidemiology, Harvard TH Chan School of Public Health, Boston, Massachusetts, United States of America

## Abstract

**Objective:** To develop and validate the Transformer-based Risk assessment survival model (TRisk), a novel deep learning model, for prediction of 10-year risk of cardiovascular disease (CVD) in both the general population and individuals with diabetes.

**Design:** Prospective open cohort study design.

**Setting:** Primary and secondary care in England as provided by Clinical Practice Research Datalink (CPRD) Gold

**Participants:** An open cohort of 3 million adults aged 25 to 84 years was identified using linked primary and secondary electronic health records from 291 and 98 general practices in England and were used for model development and validation, respectively (i.e., general population cohort). Additionally, a second cohort of patients with diabetes was extracted. At study entry, patients in both cohorts were free of CVD and not prescribed statins.

**Methods:** TRisk utilised all diagnosis, medication, procedure, and clinical test data up to study entry in linked longitudinal primary and secondary care electronic health records for prediction of 10-year risk of CVD. Discrimination, calibration, and decision curve analyses were conducted to investigate predictive performance. The proposed model was also compared against QRISK3 and a deep learning derivation model of QRISK3 (DeepSurv). Additional analyses compared discriminatory performance in other age groups, by sex, and across categories of socioeconomic status.

**Main outcome measures:** Incident cardiovascular disease recorded in either linked general practice or hospital admission datasets provided by CPRD Gold.

**Results:** TRisk demonstrated superior discrimination (C-index in the general population: 0.910; 95% confidence interval [CI]: 0.906 to 0.913). TRisk’s performance was found to be less sensitive to population age range than the benchmark models and outperformed other models also in analyses stratified by age, sex or socioeconomic status. All models were overall well-calibrated. In decision curve analyses, TRisk demonstrated greater net benefit than benchmark models across the range of relevant thresholds. At both the recommended 10% risk threshold and the 15% risk threshold, TRisk reduced both the total number of patients classified at high risk (by 22% and 35% respectively) and the number of false negatives as compared with currently recommended strategies. TRisk similarly outperformed other models in patients with diabetes. Compared with the widely recommended treat-all policy approach for patients with diabetes, TRisk at a 10% risk threshold would lead to deselection of 24% of individuals with a small fraction of false negatives (0.2% of cohort).

**Conclusion:** TRisk enabled a more targeted selection of individuals at risk of CVD compared to benchmark statistical and deep learning models, in both the general population and patients with diabetes. Incorporation of TRisk into routine clinical care would allow a reduction in the number of treatment-eligible patients by approximately one-third while preventing at least as many events as with currently adopted approaches.

## Introduction

Blood pressure (BP) and LDL-cholesterol lowering are well-established and common strategies for preventing cardiovascular diseases (CVD). The results from randomised controlled trials have shown that relying on a single risk factor is inadequate for selecting individuals for treatment, as the relative risk reductions achieved by antihypertensives and statins remain largely consistent regardless of baseline BP, LDL-cholesterol, and other factors such as age, sex, and pre-existing vascular disease.^1–4^ Therefore, it is crucial to predict an individual’s risk of CVD in order to identify those who will benefit the most from preventative treatment. Consequently, using multivariable CVD risk prediction tools has become a routine practice for clinical decision-making.^5^ This approach is strongly supported by randomised evidence of greater risk reductions for those with higher predicted CVD risk.^2,6^

Methods for identifying high-risk individuals vary globally. However, in general, guidelines for primary prevention of CVD recommend the use of risk prediction models. For example, the UK NICE guideline recommend treatment for individuals with a predicted 10-year CVD risk of 10% or more, as estimated by QRISK2.^5^ The European guidelines adopt a similar approach based on the SCORE2 model.^7^ Guidelines also often classify specific patient groups automatically as high-risk, warranting treatment without formal risk assessment. This is common for secondary prevention in patients with a history of CVD or individuals with other “high-risk” conditions such as diabetes or chronic renal disease.^7^ While these approaches are more efficient and cost-effective compared to simpler clinical criteria (e.g., based on age or BP threshold), they are still relatively crude.^8^ For instance, applying the QRISK2 model to the UK population deems approximately one-third of adults aged 30-79 years eligible for antihypertensive therapy.^9,10^ The relatively low specificity of models at the recommended thresholds means that even without treatment, most individuals who were offered treatment would not experience an event. Similar challenges exist for individuals with diabetes or other high-risk conditions who have conventionally received a blanket recommendation for treatment.^11^ For example, it has been demonstrated that the risk of CVD in diabetic patients varies substantially due to differences in screening and diagnosis methods.^12^ Current policies recommending BP and LDL-cholesterol lowering treatment for all patients with diabetes fail to account for this variability in risk.

Emerging as a potentially promising solution, the Transformer, a deep learning (DL) model initially designed for natural language processing, has become a cornerstone of applied artificial intelligence (AI) research.^13^ By being able to efficiently analyse multimodal sequential data, Transformers have shown great promise in fields such as computer vision and clinical risk prediction.^14^ Given their potential, it is crucial to fully evaluate the utility of Transformers for clinical decision-making. To this end, we conducted a study to develop and validate the Transformer-based risk assessment model (TRisk) for 10-year prediction of CVD risk.

## Methods

### Study framework

Our primary model, TRisk, was an adaptation of the Transformer-based DL model, Bidirectional EHR Transformer (BEHRT).^14^ BEHRT has demonstrated promising performance for a variety of risk prediction tasks including heart failure, stroke, and coronary heart disease.^14–16^ In this study, we extended and modified BEHRT from a binary outcome prediction model into a survival model to additionally account for censoring and refer to it as TRisk.^17^ We compared TRisk with two expert-guided models as the best-in-class models for CVD risk prediction: the QRISK3 model ^18^ and a locally derived non-linear DL derivation of Cox proportional hazards (CPH) modelling called DeepSurv, a multi-layer perceptron (MLP) neural network model.^19^ All models were validated in a general population cohort (i.e., patients without CVD at baseline).

### Data source and validation strategy

We used Clinical Practice Research Datalink (CPRD) as the study data source.^1^ CPRD includes detailed patients’ records such as demographics (age, sex, ethnicity), diagnoses, prescribed treatments, tests, and health related lifestyles collected from participating general practices (GPs) across UK, covering around 7% of the UK population. With linkage to Hospital Episode Statistics (HES) and Office for National Statistics for data about hospital admission and death registration, respectively, CPRD is one of the most comprehensive deidentified EHR datasets broadly representative of the UK population.^20^

We included data from 389 contributing GPs that met acceptable standards of research quality. Prior to cohort selection, as our validation strategy, we randomly allocated three quarters (i.e., 298) of practices to the derivation dataset and the rest (i.e., 91) of the practices to the validation dataset (details in Supplementary Methods: Clarification on validation study).^18^

### Cohort selection

We identified an open cohort for our analysis consisting of individuals without prior CVD (defined as any of coronary heart disease, stroke, and transient ischaemic attack), in whom treatment is recommended when their predicted risk of CVD is above a certain threshold.^5,7^ We included individuals who had records between 1 January 1998 and 31 December 2015, were aged between 25 and 84 years old, and registered with GP for at least one year. Following previous CVD prediction works, the index date (baseline) was randomly selected from the patient medical history for each individual.^21,22^ This method for index date selection was implemented to ensure the model was trained and evaluated on a more representative spread of age and calendar year at baseline. By adopting this approach, we simulate the calculation of a patient’s risk score at any point during the eligible period as opposed to fixing the index date as first date in eligible period, closely mirroring its real-world application in clinical settings.^21^

We aimed to adhere closely to the criteria for selection of the cohort as reported for the QRISK3 study.^18^ Similar to the QRISK3 derivation study, patients with a prior history of CVD (as defined previously) and prescription of a statin were excluded from datasets. In addition to these filtering steps, we further excluded those without information on index of multiple deprivation (IMD), without linkage to HES, and patients for whom there were no available records at the index date. We excluded the latter group as conducting inference on patients without any data for input at the index date offers little clinical value. Patients were censored at the earliest of the last date in practice, last collection from practice, death, incident CVD, 10 years after the index date (i.e., truncation after 120-month mark), or the study end date (31 December 2015).

### Outcome definition

The outcome of interest was the 10-year risk of major CVD events. CVD was defined as a composite of coronary heart disease, ischaemic stroke, or transient ischaemic attack captured in both primary and secondary care diagnostic records. We adopted the CALIBER repository for all disease phenotyping.^23^ Specifically, CALIBER phenotyping dictionaries for ischaemic stroke, transient ischaemic attack, and coronary heart disease (including myocardial infarction) were used to identify the composite CVD outcome.^23^

### Model derivation

TRisk incorporated all recorded information from the following modalities in both primary and secondary care (HES) records: 3,858 distinct diagnoses, 390 categories of medications, 1,439 laboratory tests, and 679 procedures codes. As a Transformer model, TRisk considers a patient’s medical history up to baseline as a sequence which is typically of variable length. Each unit of information from the captured modalities is mapped to the patient’s age and the health service encounter, thus providing rich temporal annotations to the sequence of records (**Figure S1**). The information captured is as recorded in the structured EHR without any imputation of missing values. Free-text data and demographics such as sex or socioeconomic status were not incorporated in TRisk. As a data-driven model, TRisk was trained on raw or minimally processed EHR (Supplementary Methods: EHR pre-processing and modelling of TRisk; **Table S1**).

### Benchmark modelling

For modelling, sex-specific QRISK3 equations were implemented using the source codes provided by ClinRisk Limited. Additionally, a non-linear DL derivation of QRISK3 was derived and validated on our general population cohort. Extraction and transformation of predictors for QRISK3^18^ followed roughly the specifications of previously published work. Variables used in the QRISK3 model were also used for DeepSurv. As a MLP neural network model, DeepSurv includes the same fixed-length cross-sectional variables as in QRISK3 and is not designed to capture variable-length longitudinal variables as TRisk does.^19^ To address data missingness, imputation was separately carried out on derivation and validation datasets. More information on predictor extraction, imputation for benchmark modelling, and implementation can be found in Supplementary Methods: Predictor selection for benchmark models and Implementation details (**Table S2**).

### Performance analysis

We analysed the performance of all models on the validation dataset using complementary approaches. Model discrimination was assessed by calculating the concordance index (C-index) and area under the precision-recall curve (AUPRC). While C-index is a survival risk prediction metric that evaluates the model’s ability to rank order cases and non-cases without consideration of event rate, AUPRC is a prediction performance metric that implicitly takes into account event rate into its formulation; specifically, a model making random predictions would exhibit an AUPRC equal to the event rate and a C-index of 0.5. AUPRC summarizes the trade-off between positive predictive value and sensitivity for different thresholds proving especially useful in cases of imbalanced label prediction, such as incident CVD prediction. Consistency of predictions at the level of individuals was assessed with a scatterplot in order to investigate the agreement between the models. Decision curve analysis (accounting for censoring) was conducted to plot the trade-off between correctly captured CVD cases (true positives) and incorrectly captured high-risk individuals without a CVD event (false positives or “false alarms”) across the spectrum of reasonable risk thresholds.^24^ We further analysed the impact of TRisk for preventative therapy recommendation by calculating the number of patients who would be classified as high risk, the number of true positives, and the number of false negatives at specific illustrative thresholds. Censoring was not accounted for in this analysis. While the selection of an optimal threshold for decision-making cannot be reduced to performance metrics only, a superior strategy would maximise capture of those who are in need of preventative therapy (i.e., minimising false negatives) whilst minimising false alarms. For all aforementioned analyses across all models, in order to derive patient-level risk, we estimated the survival function over the follow-up period with a specific focus on the risk estimates at the 10^th^ year.^18^ While for some models (e.g., QRISK3), validation has been conducted separately on male and female patients, we have presented results utilising the aggregate set of predictions on all patients in the validation dataset.

In accordance with the prior QRISK3 study, our main analyses were restricted to patients aged 25 to 84 years.^18^ However, in additional analyses, we assessed model discrimination in terms of C-index in the age range of 40 to 69 years and 40 to 84 years. Model discrimination was further assessed separately in men and women, and at different levels of socioeconomic deprivation denoted by quantiles of IMD.

Furthermore, for additional comparison against TRisk, we derived and validated a sex-agnostic CPH model with the same predictor set as the established, SCORE2 model.^25^ More details on predictor extraction, imputation, and implementation of this locally derived CPH model can be found in Supplementary Methods: Predictor selection for benchmark models and Implementation details.

Lastly, we repeated all aforementioned analyses on patients with a diagnosis of diabetes prior to or on the index date (i.e., diabetes cohort) to assess the usefulness of TRisk in a ‘high-risk’ population. By utilising a transfer learning approach, the proposed TRisk model that was initially trained on the general population cohort was transferred, fine-tuned, and validated in patients with diabetes at baseline (Supplementary Methods: Modelling in diabetes cohort).^26^

The approval for this work was given by the CPRD Independent Scientific Advisory Committee of UK (protocol number: 16_049R).

### Patient involvement

Patients were not involved in the design and conduct of this research.

## Results

2.97 million patients were included in the general population cohort (747,076 patients in validation) median follow-up of 2.5 (interquartile range [IQR]: [0.8, 5.9]) years. 4.6% of patients were diagnosed with CVD during follow-up (4.7% in derivation; 4.5% in validation) (**Table 1**). As expected, there was a noticeable variation in the distribution of regions between the derivation and validation datasets in the cohort.

**Table 1.** Population characteristics for derivation and validation datasets of general population cohort.

|  | Derivation<br>(n=2,224,701) | Validation<br>(n=747,076) |
| --- | --- | --- |
| CVD cases (%) | 104,058 (4.68) | 33,248 (4.45) |
| Women (%) | 1,202,114 (54.03) | 402,962 (53.94) |
| Mean age in years (SD) | 48 (16) | 47 (16) |
| Ethnicity |  |  |
| Unknown (%) | 1,481,771 (66.61) | 498,763 (66.76) |
| White (%) | 691,797 (31.1) | 231,088 (30.93) |
| Other Asian (%) | 5,399 (0.24) | 1,804 (0.24) |
| Pakistani (%) | 5,673 (0.26) | 1,358 (0.18) |
| Indian (%) | 9,707 (0.44) | 2,528 (0.34) |
| Other (%) | 9,324 (0.42) | 3,492 (0.47) |
| Caribbean (%) | 4,187 (0.19) | 2,154 (0.29) |
| Mixed (%) | 3,615 (0.16) | 1,396 (0.19) |
| Bangladeshi (%) | 1,707 (0.08) | 449 (0.06) |
| Chinese (%) | 2,352 (0.11) | 805 (0.11) |
| Other Black (%) | 2,715 (0.12) | 1,021 (0.14) |
| Black African (%) | 6,454 (0.29) | 2,218 (0.30) |
| Index of multiple deprivation (IMD) |  |  |
| IMD 1 (%) | 526,997 (23.69) | 168,810 (22.60) |
| IMD 2 (%) | 500,928 (22.52) | 155,571 (20.82) |
| IMD 3 (%) | 454,225 (20.42) | 167,026 (22.36) |
| IMD 4 (%) | 396,469 (17.82) | 147,640 (19.76) |
| IMD 5 (%) | 346,082 (15.56) | 108,029 (14.46) |
| Region |  |  |
| North East (%) | 31,767 (1.43) | 26,764 (3.58) |
| North West (%) | 331,324 (14.89) | 68,397 (9.16) |
| Yorkshire and the Humber (%) | 78,998 (3.55) | 42,732 (5.72) |
| East Midlands (%) | 60,070 (2.70) | 37,876 (5.07) |
| West Midlands (%) | 247,430 (11.12) | 92,491 (12.38) |
| East of England (%) | 271,342 (12.2) | 84,811 (11.35) |
| South West (%) | 239,212 (10.75) | 123,501 (16.53) |
| South Central (%) | 322,056 (14.48) | 55,236 (7.39) |
| London (%) | 363,896 (16.36) | 115,077 (15.4) |
| South East Coast (%) | 278,606 (12.52) | 100,191 (13.41) |
| Mean systolic blood pressure<sup>†</sup>, mmHg (SD) | 129.13 (14.04) | 128.77 (14.07) |
| Mean body mass index<sup>†</sup>, kg/m<sup>2</sup> (SD) | 27.0 (3.89) | 26.93 (3.90) |
| Mean high density lipoprotein<sup>†</sup>, mmol/L (SD) | 1.39 (0.66) | 1.34 (0.70) |
| Mean total cholesterol<sup>†</sup>, mmol/L (SD)<sup>†</sup> | 4.93 (3.15) | 5.08 (3.96) |
| Smoking status<sup>†</sup> (number of cigarettes per day) |  |  |
| Non-smoker (%) | 1,145,267 (51.48) | 388,858 (52.05) |
| Ex-smoker (%) | 580,394 (26.09) | 187,931 (25.16) |
| Light (<10 cigarettes/day) (%) | 145,747 (6.55) | 51,860 (6.94) |
| Moderate (10-20 cigarettes/day) (%) | 211,134 (9.49) | 71,394 (9.56) |
| Heavy (>20 cigarettes/day) (%) | 142,159 (6.39) | 47,033 (6.30) |
| Comorbidities at baseline |  |  |
| Diabetes (%) | 44,668 (2.01) | 14,518 (1.94) |
| Rheumatoid arthritis (%) | 10,838 (0.49) | 3,602 (0.48) |
| Atrial fibrillation (%) | 26,889 (1.21) | 8,565 (1.15) |
| CKD (%) | 7,351 (0.33) | 2,470 (0.33) |
| Migraine (%) | 78,559 (3.53) | 26,263 (3.52) |
| Lupus erythematosus (%) | 1,755 (0.08) | 557 (0.07) |
| Mental illness (%) | 17,007 (0.76) | 6,223 (0.83) |
| HIV/AIDS (%) | 4,139 (0.19) | 1,484 (0.20) |
| Erectile dysfunction (%) | 44,744 (2.01) | 15,386 (2.06) |

| Medication use at baseline |  |  |
| --- | --- | --- |
| Antihypertensives (%) | 99,411 (4.47) | 31,039 (4.15) |
| Antipsychotics (%) | 4,222 (0.19) | 1,374 (0.18) |
| Corticosteroids (%) | 29,362 (1.32) | 9,207 (1.23) |

SD: standard deviation; %: percentage; CVD: cardiovascular disease; IMD: Index of multiple deprivation; CKD: chronic kidney disease; <sup>†</sup>: missing observations: smoking status (48% missing in general population cohort), systolic blood pressure (39%), standard deviation of systolic blood pressure (64%), body mass index (59%), total cholesterol (73%), and high-density lipoprotein cholesterol (81%).

### Model performance analyses on validation data

In terms of discrimination, TRisk demonstrated higher C-index and AUPRC as compared to QRISK3 and DeepSurv models (**Table 2; Table S3)**. Assessing calibration, while QRISK3 showed some deviation from perfect calibration, all models examined in this study generally exhibited acceptable calibration within the typically relevant threshold range (i.e., 0-20%) for decision making (**Figure 1A; Figure S2**). Comparison of the predictive distribution of the models (**Figure 1B**) showed that the benchmark models had a narrower distribution of predicted risk than TRisk. By contrast, TRisk showed a wider distribution with classification of higher fraction of patients to the very high and very low risk ranges. Comparing the consistency of individual-level predictions, with QRISK3 as reference model, we found that while benchmark models were largely ranking patients consistently among one other, there was poor correlation with TRisk, which was better at identifying true positive cases (**Figure S3**).

**Figure 1.**
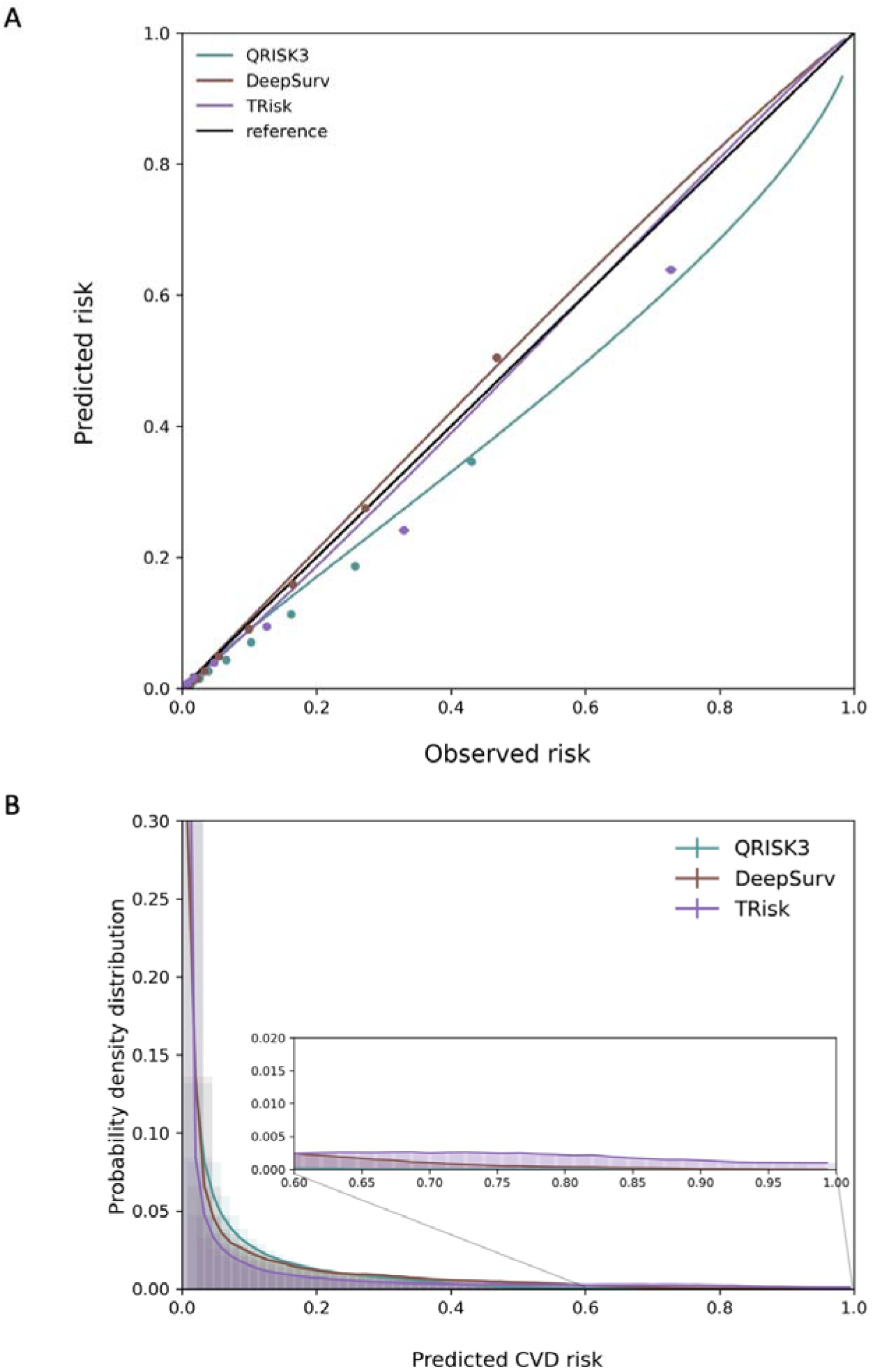
Calibration plots and distribution of predicted risk of models. Smoothed calibration lines and tenth of predicted risk decile calibration plots (A) and distribution of predicted risk (B) are presented for models implemented on general population cohort.

**Table 2.** Discrimination performance of models in general population cohort.

| Model | Concordance-index (95% CI) |
| --- | --- |
| QRISK3 | 0.831 (0.826, 0.835) |
| DeepSurv | 0.846 (0.841, 0.850) |
| TRisk | 0.910 (0.906, 0.913) |

*CI: confidence interval*

In analyses investigating populations in different age bands, the gap in discrimination performance between TRisk and the benchmark models were found to become larger as the age range was narrowed (**Table S4)**. TRisk had a higher discrimination across all age categories. Although TRisk did not include sex and IMD as predictors, its discriminatory performance in subgroups by sex and IMD was better than the benchmark models with no significant difference between stratifications (**Tables S5, S6**).

Decision curve analysis demonstrated that across relevant thresholds for decision making, TRisk provided greater net benefit than benchmark models (**Figure 2**). In supplementary modelling analysis, we found that our locally derived CPH risk model with SCORE2 predictors performed similarly to other benchmark models across all analyses (**Figures S3-S6; Tables S3-S6**).

**Figure 2.**
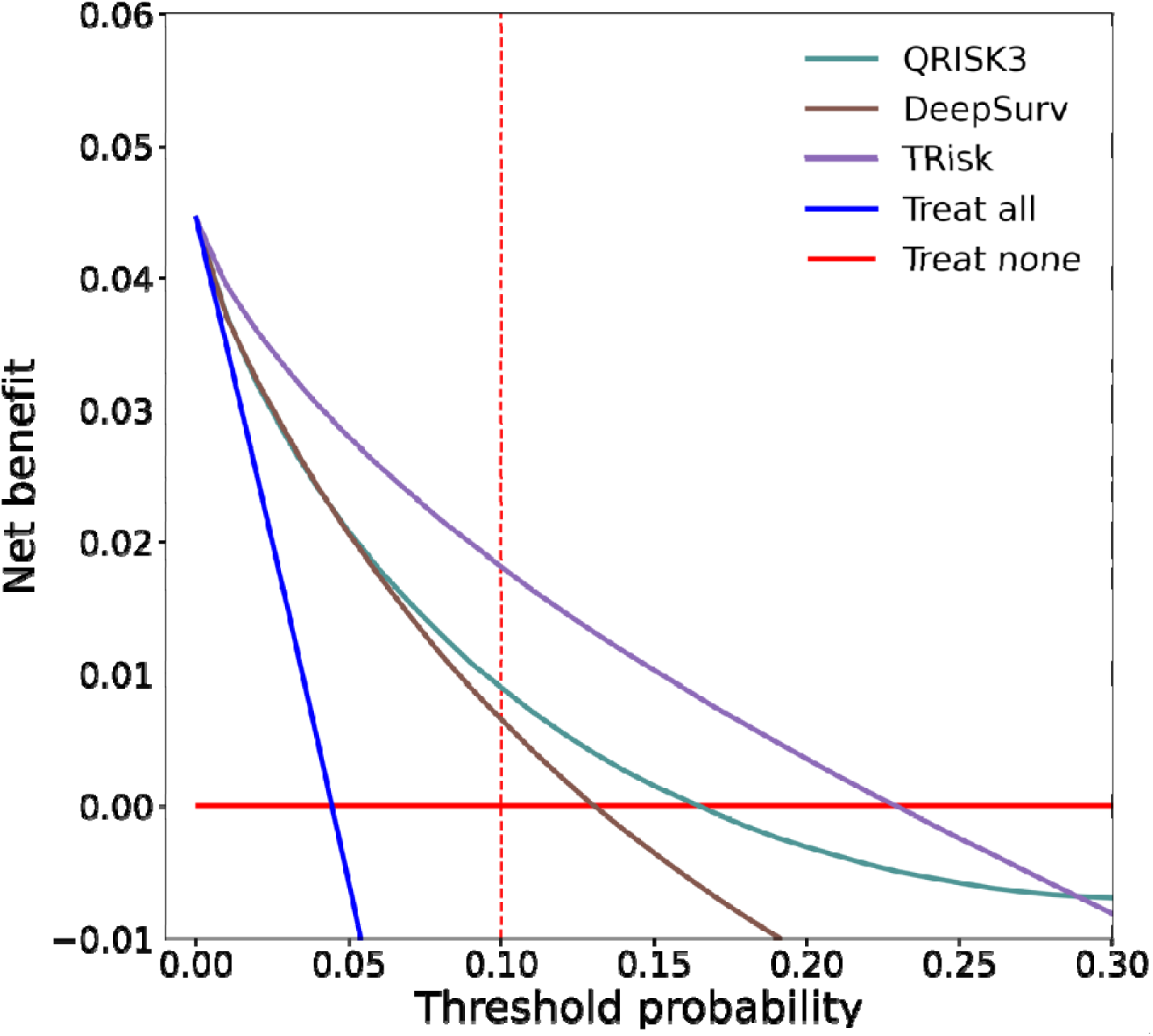
Decision curve analysis for analysed models. Decision curve analysis (including censored observations) has been conducted for models in general population cohort. 10% decision threshold used by various clinical guidelines for preventative treatment recommendation in general population is illustrated with dotted red line. Threshold probability is shown on the x-axis and the net benefit, a function of threshold probability, is shown on the y-axis and is the difference between the proportion of true positives and false positives weighted by odds of the respective decision threshold.

In analysis of the diabetes cohort, 59,186 patients with diabetes (14,518 patients in validation) were identified. Over a median follow-up of 2.3 years (IQR: [0.9, 5.0]), 12.5% suffered a CVD event (12.8% in derivation; 11.7% in validation) **(Table S7)** Model performance metrics observed in the diabetes cohort were overall concordant with those in the general population cohort (**Figures S7-S10; Tables S8-S11**).

The clinical impact analysis of risk assessment approaches at different risk thresholds standardised to a population of 1,000 patients is shown in **Table 3** (select strategies in **Figure S11**; analysis on non-standardised cohorts in **Supplementary Results**).

**Table 3.** Comparison of the clinical impact of models on selected outcomes at different risk thresholds, standardised to 1,000 patients in general population and diabetes cohorts.

| Cohort | Strategy (model) | Number of people classified as high risk | Number of events in people at high risk | Number of events in people at low risk |
| --- | --- | --- | --- | --- |
| General population | Treat all | 1000 | 45 | 0 |
|  | QRISK3 at 10% threshold (recommended) | 272 | 36 | 9 |
|  | TRisk at 10% threshold | 216 | 39 | 5 |
|  | QRISK3 at 15% threshold | 187 | 29 | 15 |
|  | TRisk at 15% threshold | 178 | 37 | 8 |
|  | QRISK3 at 20% threshold | 131 | 24 | 21 |
|  | TRisk at 20% threshold | 152 | 35 | 10 |
|  | Treat none | 0 | 0 | 45 |
| Diabetes | Treat all (recommended) | 1000 | 117 | 0 |
|  | QRISK3 at 10% threshold | 866 | 114 | 3 |
|  | TRisk at 10% threshold | 757 | 115 | 2 |
|  | QRISK3 at 15% threshold | 764 | 110 | 7 |
|  | TRisk at 15% threshold | 682 | 112 | 5 |
|  | QRISK3 at 20% threshold | 649 | 103 | 14 |
|  | TRisk at 20% threshold | 615 | 109 | 8 |
|  | Treat none | 0 | 0 | 117 |

In the general population cohort, QRISK3 at the recommended 10% risk threshold ^5,7^ would classify 272 (for every 1,000 individuals) as eligible for treatment with 36 patients suffering an event (13% true positive) over the 10-year follow-up (**Table 3; Figure S11A**). The remaining 728 patients were ranked as low-risk of whom, 720 (99%) were true negatives. Operating at 15% threshold would restrict the cohort of eligible patients for preventative therapy to 187 patients, but simultaneously increase false negatives from 9 to 15 patients (66% increase).

In comparison, at the 10% threshold, the TRisk model enabled superior true positive (and minimal false negative) capture whilst decreasing the treatment eligible population by 56 patients (21% reduction) compared with the recommended strategy (i.e., QRISK3 at 10%). Up to a threshold of 15% risk, the TRisk model effectively maintained true case capture with respect to the recommended strategy (i.e., same true positive/false negative counts) whilst reducing the number of those considered high-risk from 272 to 178 patients (35% reduction; **Table 3; Figure S11B**). At both 10% and 15% thresholds, TRisk improved upon the recommended strategy: in the former, by delivering superior true positive capture and in the latter, by enabling more selective capture of those considered high-risk without attrition in true positive/false negative capture with respect to recommended strategy.

For the diabetes cohort, the TRisk approach similarly demonstrated better positive and negative capture as compared to conventional approaches (**Table 3; Figure S11C**). At the threshold of 10%, TRisk captured a more refined population of 757 patients with 114 patients as true positives (15% true positive). As compared to the conventional indiscriminate approach of recommending all patients with diabetes for treatment, TRisk would recommend 243 fewer patients (24% reduction) with minimal false negative capture (2 per 1000 patients; 0.2% of cohort).

Recently, revised UK guidelines have recommended utilising QRISK3 as a risk assessment strategy for even those with diabetes (**Table 3; Figure S11D**).^5^ Our impact analysis yielded that QRISK3 at the recommended 10% threshold selected 866 patients for preventative therapy, with 114 true positive cases (13% true positives) with natural degradation in true positive capture at higher thresholds. In comparison, TRisk at the same threshold would recommend 109 fewer patients (13% reduction) whilst gaining one true positive patient capture (**Table 3; Figure S11E**).

## Discussion

Our study shows that TRisk, a novel Transformer-based survival model, significantly outperformed widely recommended benchmark models in identifying individuals at risk of CVD. TRisk was also less dependent on age than our benchmark models. Due to its higher discriminatory performance, TRisk enabled a potential upwards shift in the commonly recommended risk thresholds for more targeted selection of individuals for treatment. Application of TRisk would lead to selection of about one-third and one-fourth fewer individuals in general and diabetes populations respectively than application of existing policies without any material trade-offs.

Risk-based selection of individuals for CVD preventative therapy is widely used in clinical practice.^5^ However, existing approaches result in a large number of individuals being eligible for treatment, even though many of them will not experience an event.^10^ Indeed, in a survey of general physicians in the Netherlands, a key barrier to uptake of CVD risk prediction models was their potential for over-treatment.^27^ In line with this, we found QRISK3 to classify over a quarter of the adult population as eligible for treatment in the general population^10^; however, seven out of eight individuals predicted to have a preventable CVD event did not experience such an event. We found that increasing the risk threshold for such models would not overcome their prediction inefficiency; while the proportion of true positives would increase, number of false negatives would also increase, due to their relatively low sensitivity.

TRisk has the potential to lead to substantial efficiency gains without a trade-off in increasing false negative capture. At the traditional 10% risk threshold but also a higher 15% threshold, it reduced both the overall number of patients classified as high risk and the number of patients classified as being at low risk who subsequently experienced an event (i.e., false negatives). Significant performance improvements were also found for risk stratification in patients with diabetes. While the guidelines have typically recommended preventative therapy for all patients with diabetes, our analyses demonstrated that application of TRisk will enable a 24% reduction in the number of patients assigned to treatment without increasing false negatives, if treatment efficiency is a priority.

More recently, in light of evidence highlighting QRISK3’s modest but acceptable performance in those with type 2 diabetes, the UK NICE guidelines have shifted to a risk assessment strategy utilising QRISK3 at 10% for preventative treatment recommendation. Similar policies might also get adopted by other nations and societies, in order to reduce the number of treatment-eligible patients and make preventive treatments more acceptable to patients and doctors. In this study, we found that, replacing QRISK3 with TRisk would rank 13% fewer patients as above the 10% risk threshold whilst capturing more true positives.^11^

Naturally, while low costs and high safety of preventive therapy have encouraged more liberal treatment recommendation policies, there have been calls for better matching of individuals to treatments.^28^ TRisk offers one tangible solution to this challenge today. Deployment and use of TRisk could help offer medical treatment to truly high-risk individuals, whilst directing others to non-pharmacological forms of therapy (diet alterations, exercise, etc.). In the UK context, it would translate into offering treatment to 35% fewer patients while still preventing at least as many events as the QRISK3 model. However, this is not to say that preventive treatments are to be withheld from lower risk individuals. In settings, where treatment costs are low (and harms not excessive), a higher fraction of the population could be offered treatment. Our study show that even in such scenarios, TRisk is more effective and efficient in targeting individuals. Importantly, this optimised patient classification is achieved with the use of readily available information in patient records without the need for collection of additional information, for instance with imaging or biomarkers.

A key concern with AI in medicine has been its potential to bias against particular patient groups.^29^ In this study, we found no such evidence for TRisk at least in major patient subgroups. In analyses stratified by sex and socioeconomic status, TRisk showed higher discriminatory performance and lower variation than all benchmark models even though these features were not explicitly included in the TRisk. Of note, we found that TRisk was less reliant on age than the benchmark models. As the age range was narrowed, the performance gap between TRisk and benchmark models widened. Age is typically a proxy of unmeasured or poorly measured risk factors and by utilising the rich latent features captured from temporal, minimally processed EHR, the reliance on age as a proxy diminishes. By contrast, the discriminatory performance of all benchmark modelling approaches diminished substantially when the same age groups were applied to them.^18,25^ Indeed, within the same age groups, the performance of these traditional models was very similar.

How are Transformer-based models like TRisk able to make predictions, outperforming expert-driven models? The input space of TRisk includes all recorded diagnoses, medications, laboratory measurements, and procedures in a patient’s primary care or hospital records. TRisk captures the presence or absence of such information in their temporal context to predict the next event. The multiple layers of abstraction of such information limits a simplistic identification of individual predictors that are likely to ‘independently’ apply to all patients and across all times. However, previous explainability analysis has shown that EHR Transformers can implicitly capture patient sex despite not including it as a variable and even identify epidemiologically known risk factors while capturing associations that are less well-known or typically not considered.^14,15^ For instance, iron deficiency anaemia or use of treatments for chronic obstructive pulmonary disease are not typically considered as risk factors for heart failure but were identified by the model.^15^ It was also able to detect associations that varied over time. For instance, association of drugs used for the treatment of glaucoma with heart failure showed a gradual quantitative and qualitative shift over years, which corresponded to a change in glaucoma treatment from beta-blockers to prostaglandin analogues over time.^15^ It seems likely that by adding more data modalities such as omics (which TRisk can accommodate), we will see further improvements in predictions, particularly in special patient subgroups.

In terms of benchmark modelling, in our implementation of QRISK3, we found that despite some differences in methodologies, our findings of QRISK3 performance were broadly in line with those from the derivation study.^18^ Notably, unlike the derivation study, we did not have access to Townsend scores for modelling, excluded patients who did not have any records, implemented the random sampling method for selection of baseline in eligible period, and used codes published by the CALIBER group to identify the CVD outcome.^23^ Due to potentially these differences, we observed both, higher incident CVD event rate of 4.6% and mean age of 47 years with respect to figures from the original study.^18^ Nevertheless, discrimination (C-index: ~0.83) findings were consistent with those reported in the original validation study and other independent analyses (approximately 0.84-0.88).^18,21,30^ Although we found slight deviation from perfect calibration, our implementation of QRISK3 was generally found to be acceptably calibrated in the range of interest (i.e., up to 20% decision boundary). In gist, we found that our implementation of QRISK3 performed similarly to previously published reports.

Indeed, while such models such as QRISK3 or the SCORE class of models have demonstrated strong discrimination performance in primary prevention population studies, the same models for patients with pre-existing conditions such as diabetes are less explored and tend to face performance degradation.^31^ Found in published evidence and in our independent analyses, the performance attenuation might be due to the differing relationship of established risk factors with each other in addition to metabolic specific risk factors and the outcomes that is not entirely consistent with the knowledge in the general population.^31,32^ Utilising TRisk as a conduit for knowledge transfer across cohorts resulted in strong predictive performance for the diabetes cohort. Specifically, the superior performance was made possible by initially learning risk patterns from a more homogeneous and significantly larger general population cohort. The relationships and weights obtained from this initial learning phase are then applied to the smaller and more complex subgroup of diabetic patients and fine-tuned, rather than training the model from randomly initialized weights.^26^

This approach not only improves risk assessment performance within a specific subgroup but also moves towards a unified data-driven model for predicting CVD risk, eliminating the need for separate models in different patient subgroups. The successful transfer of knowledge from the general population to subgroups in the case of cardiovascular risk prediction suggests that the potential of such “group to subgroup” knowledge transfer should be further explored in various clinical contexts.^26^

### Strengths and limitations

In this study, we have conducted comprehensive multi-dimensional comparisons of model types (statistical and DL) applied to two different target groups (general risk and high risk) by using multiple metrics of evaluation: discrimination, calibration, net benefit, and impact analyses at different risk thresholds.

There are some notable aspects of our study that illuminate important focus areas for future research. Indeed, the exclusion of statin users at baseline has been shown to increase bias in prediction as patients may start statins soon after baseline.^33^ Hence, while it is appreciated that exclusion of statins might lead to underestimation of risk, in this work, we intended to conduct prediction in cohort free of statin-use as conducted by the QRISK3 derivation study. Future research should investigate approaches that can potentially account for “treatment drop-in.”

In terms of transportability of TRisk, further evaluation of TRisk’s adaptability across various data landscapes, additional patient subgroups, and clinical outcomes is necessary. Future research should actively explore TRisk in other UK EHR data settings (e.g., using data from other UK EHR systems) and other international data settings as well. Nevertheless, a few features of the study are of note and provide some preliminary assurances on wider generalisability.^34^ Although TRisk was validated on one of the representative UK EHR datasets (akin to other widely accepted benchmark approaches such as QRISK3^18^), it was deliberately engineered to be transferable across various data settings. Specifically, TRisk was designed to theoretically operate on routinely collected data standardised through established clinical dictionaries and mapping libraries (e.g., SNOMED CT) which streamline the translation of disease, medication, procedure, and other clinical codes across different healthcare systems. While this fundamental design feature enhances TRisk’s chances of adaptability, downstream studies in various EHR data settings are necessary to fully evaluate the transportability of the proposed model.

Lastly, as another angle of transportability and validation, TRisk should also be evaluated on other outcomes in addition to CVD. Certainly, previous research has demonstrated that TRisk’s foundational BEHRT model surpasses several other approaches in multi-outcome prediction of over 300 clinical outcomes, hence providing initial evidence that TRisk can serve as a multipurpose risk assessment tool.^14^ Complementarily to this body of evidence, several research groups have independently applied BEHRT and variations in other EHR systems across the USA (e.g., New York, Florida, California) for a host of clinical outcome prediction tasks (e.g., emergency admission, COVID-19 related outcomes) across diverse patients groups – ultimately, providing strong, promising evidence that underscores the potential wide-ranging adaptability of the TRisk approach.^35–37^ TRisk depends on access to the entire EHR of an individual and cannot be reduced to a simple scoring algorithm. While this adds further complexity to implementation of such models into practice compared with simpler regression models, there are effective tools that have been robustly tested to facilitate the transition of AI from development to deployment in settings with limited computational capacities.^38^ Indeed, future efforts could explore leveraging TRisk as a first-line screening tool for population-level risk assessment alerting GPs of potential at-risk patients that require further clinical attention.

## Conclusion

TRisk, a novel Transformer-based survival model, outperformed standard models and recommendations for selection of individuals at high risk of CVD. Implementation of TRisk into routine practice could improve allocative efficiency by reducing the number of patients offered treatment by about one-third and one-fourth fewer individuals in general and diabetes populations respectively as compared with status-quo recommendation strategies.

## Data Availability

All coding libraries produced in the present study are available upon reasonable request to the authors

## References

1. The Blood Pressure Lowering Treatment Trialists’ Collaboration. Pharmacological blood pressure lowering for primary and secondary prevention of cardiovascular disease across different levels of blood pressure: an individual participant-level data meta-analysis. The Lancet 2021; 397: 1625–1636.

2. Mihaylova B, Emberson J, Blackwell L, Keech A, Simes J, Barnes EH, Voysey M, Gray A, Collins R, Baigent C, De Lemos J, Braunwald E, Blazing M, Murphy S, Downs JR, Gotto A, Clearfield M, Holdaas H, Gordon D, et al. The effects of lowering LDL cholesterol with statin therapy in people at low risk of vascular disease: Meta-analysis of individual data from 27 randomised trials. The Lancet 2012; 380: 581–590.

3. Bidel Z, Nazarzadeh M, Canoy D, Copland E, Gerdts E, Woodward M, Gupta AK, Reid CM, Cushman WC, Wachtell K, Teo K, Davis BR, Chalmers J, Pepine CJ, Rahimi K. Sex-Specific Effects of Blood Pressure Lowering Pharmacotherapy for the Prevention of Cardiovascular Disease: An Individual Participant-Level Data Meta-Analysis. Hypertension. Epub ahead of print 24 July 2023. DOI: 10.1161/HYPERTENSIONAHA.123.21496.

4. Fulcher J, O’Connell R, Voysey M, Emberson J, Blackwell L, Mihaylova B, Simes J, Collins R, Kirby A, Colhoun H, Braunwald E, La Rosa J, Pedersen TR, Tonkin A, Davis B, Sleight P, Franzosi MG, Baigent C, Keech A, et al. Efficacy and safety of LDL-lowering therapy among men and women: Meta-analysis of individual data from 174 000 participants in 27 randomised trials. The Lancet; 385. Epub ahead of print 2015. DOI: 10.1016/S0140-6736(14)61368-4.

5. Nat’l Inst for Health and Care Excellence. *Lipid modification: cardiovascular risk assessment and the modification of blood lipids for the primary and secondary prevention of cardiovascular disease | introduction | Guidance and guidelines | NICE*. 2014.

6. Sundström J, Arima H, Woodward M, Jackson R, Karmali K, Lloyd-Jones D, Baigent C, Emberson J, Rahimi K, Macmahon S, Patel A, Perkovic V, Turnbull F, Neal B, Agodoa L, Estacio R, Schrier R, Lubsen J, Chalmers J, et al. Blood pressure-lowering treatment based on cardiovascular risk: A meta-analysis of individual patient data. The Lancet 2014; 384: 591–598.

7. Visseren FLJ, Mach F, Smulders YM, Carballo D, Koskinas KC, Back M, Benetos A, Biffi A, Boavida JM, Capodanno D, Cosyns B, Crawford C, Davos CH, Desormais I, Di Angelantonio E, Franco OH, Halvorsen S, Richard Hobbs FD, Hollander M, et al. 2021 ESC Guidelines on cardiovascular disease prevention in clinical practice. European Journal of Preventive Cardiology; 29. Epub ahead of print 2022. DOI: 10.1093/eurjpc/zwab154.

8. Karmali K, Lloyd-Jones D, Leeuw J van der, al. et. Blood pressure-lowering treatment strategies based on cardiovascular risk versus blood pressure: a meta-analysis of individual participant data. PLoS Med; 15.

9. Hippisley-Cox J, Coupland C, Vinogradova Y, Robson J, Minhas R, Sheikh A, Brindle P. Predicting cardiovascular risk in England and Wales: Prospective derivation and validation of QRISK2. BMJ 2008; 336: 1475–1482.

10. Herrett E, Gadd S, Jackson R, al. et. Eligibility and subsequent burden of cardiovascular disease of four strategies for blood pressure-lowering treatment: a retrospective cohort study. Lancet 2019; 394: 663–671.

11. NICE UK. Type 2 Diabetes in adults: management. NICE guideline. NICE guideline.

12. Pylypchuk R, Wells S, Kerr A, Poppe K, Harwood M, Mehta S, Grey C, Wu BP, Selak V, Drury PL, Chan WC, Orr-Walker B, Murphy R, Mann J, Krebs JD, Zhao J, Jackson R. Cardiovascular risk prediction in type 2 diabetes before and after widespread screening: a derivation and validation study. The Lancet; 397. Epub ahead of print 2021. DOI: 10.1016/S0140-6736(21)00572-9.

13. Vaswani A, Shazeer N, Parmar N, Uszkoreit J, Jones L, Gomez AN, Kaiser L, Polosukhin I. Attention Is All You Need. Adv Neural Inf Process Syst.

14. Li Y, Rao S, Solares JRA, Hassaine A, Ramakrishnan R, Canoy D, Zhu Y, Rahimi K, Salimi-Khorshidi G. BEHRT: Transformer for Electronic Health Records. Sci Rep 2020; 10: 7155.

15. Rao S, Li Y, Ramakrishnan R, Hassaine A, Canoy D, Cleland JG, Lukasiewicz T, Salimi-Khorshidi G, Rahimi K. An explainable Transformer-based deep learning model for the prediction of incident heart failure. IEEE J Biomed Health Inform 2022; 1–1.

16. Li Y, Salimi-Khorshidi G, Rao S, Canoy D, Hassaine A, Lukasiewicz T, Rahimi K, Mamouei M. Validation of risk prediction models applied to longitudinal electronic health record data for the prediction of major cardiovascular events in the presence of data shifts. European Heart Journal - Digital Health 2022; ztac061.

17. Tang W, Ma J, Mei Q, Zhu J. SODEN: A Scalable Continuous-Time Survival Model through Ordinary Differential Equation Networks, http://arxiv.org/abs/2008.08637 (2020).

18. Hippisley-Cox J, Coupland C, Brindle P. Development and validation of QRISK3 risk prediction algorithms to estimate future risk of cardiovascular disease: Prospective cohort study. BMJ; 357.

19. Katzman JL, Shaham U, Cloninger A, Bates J, Jiang T, Kluger Y. DeepSurv: Personalized treatment recommender system using a Cox proportional hazards deep neural network. BMC Med Res Methodol; 18.

20. Herrett E, Gallagher AM, Bhaskaran K, Forbes H, Mathur R, Staa T van, Smeeth L. Data Resource Profile: Clinical Practice Research Datalink (CPRD). Int J Epidemiol 2015; 44: 827–836.

21. Li Y, Sperrin M, Ashcroft DM, van Staa TP. Consistency of variety of machine learning and statistical models in predicting clinical risks of individual patients: Longitudinal cohort study using cardiovascular disease as exemplar. The BMJ; 371. Epub ahead of print 2020. DOI: 10.1136/bmj.m3919.

22. Van Staa TP, Gulliford M, Ng ESW, Goldacre B, Smeeth L. Prediction of cardiovascular risk using framingham, ASSIGN and QRISK2: How well do they predict individual rather than population risk? PLoS One; 9. Epub ahead of print 2014. DOI: 10.1371/journal.pone.0106455.

23. Kuan V, Denaxas S, Gonzalez-Izquierdo A, Direk K, Bhatti O, Husain S, Sutaria S, Hingorani M, Nitsch D, Parisinos CA, Lumbers RT, Mathur R, Sofat R, Casas JP, Wong ICK, Hemingway H, Hingorani AD. A chronological map of 308 physical and mental health conditions from 4 million individuals in the English National Health Service. Lancet Digit Health 2019; 1: e63–e77.

24. Vickers AJ, Elkin EB. Decision curve analysis: a novel method for evaluating prediction models. Med Decis Making 2006; 26: 565–574.

25. SCORE2 risk prediction algorithms: New models to estimate 10-year risk of cardiovascular disease in Europe. Eur Heart J; 42. Epub ahead of print 2021. DOI: 10.1093/eurheartj/ehab309.

26. Zhuang F, Qi Z, Duan K, Xi D, Zhu Y, Zhu H, Xiong H, He Q. A Comprehensive Survey on Transfer Learning, http://arxiv.org/abs/1911.02685 (2019).

27. Eichler K, Zoller M, Tschudi P, Steurer J. Barriers to apply cardiovascular prediction rules in primary care: a postal survey. BMC Fam Pract 2007; 8: 1.

28. The Lancet. Personalised medicine in the UK. The Lancet 2018; 391: e1.

29. Obermeyer Z, Topol EJ. Artificial intelligence, bias, and patients’ perspectives. The Lancet 2021; 397: 2038.

30. Livingstone S, Morales DR, Donnan PT, Payne K, Thompson AJ, Youn JH, Guthrie B. Effect of competing mortality risks on predictive performance of the QRISK3 cardiovascular risk prediction tool in older people and those with comorbidity: external validation population cohort study. Lancet Healthy Longev; 2. Epub ahead of print 2021. DOI: 10.1016/S2666-7568(21)00088-X.

31. Dziopa K, Asselbergs FW, Gratton J, Chaturvedi N, Schmidt AF. Cardiovascular risk prediction in type 2 diabetes: a comparison of 22 risk scores in primary care settings. Diabetologia; 65. Epub ahead of print 2022. DOI: 10.1007/s00125-021-05640-y.

32. Rao S, Li Y, Nazarzadeh M, Canoy D, Mamouei M, Hassaine A, Salimi-Khorshidi G, Rahimi K. Systolic Blood Pressure and Cardiovascular Risk in Patients with Diabetes: A Prospective Cohort Study. Hypertension 2023; 80: 598–607.

33. Niels Peek MSMMT van SIB. Hari Seldon, QRISK3, and the Prediction Paradox. BMJ.

34. Reilly BM, Evans AT. Translating clinical research into clinical practice: Impact of using prediction rules to make decisions. Annals of Internal Medicine; 144. Epub ahead of print 2006. DOI: 10.7326/0003-4819-144-3-200602070-00009.

35. Huang D, Cogill S, Hsia RY, Yang S, Kim D. Development and external validation of a pretrained deep learning model for the prediction of non-accidental trauma. NPJ Digit Med; 6. Epub ahead of print 2023. DOI: 10.1038/s41746-023-00875-y.

36. Pang C, Jiang X, Kalluri KS, Spotnitz M, Chen R, Perotte A, Natarajan K. CEHR-BERT: Incorporating temporal information from structured EHR data to improve prediction tasks. Proceedings of Machine Learning Research 2021; 158: 239–260.

37. Rupp M, Peter O, Pattipaka T. ExBEHRT: Extended Transformer for Electronic Health Records. 2023. Epub ahead of print 2023. DOI: 10.1007/978-3-031-39539-0_7.

38. Akter S, Michael K, Uddin MR, McCarthy G, Rahman M. Transforming business using digital innovations: the application of AI, blockchain, cloud and data analytics. Ann Oper Res; 308. Epub ahead of print 2022. DOI: 10.1007/s10479-020-03620-w.

