## Supplementary for "Refined selection of individuals for preventive cardiovascular disease treatment with a Transformer-based risk model"

### Supplementary Methods

#### Clarification on validation study

Patients from different GPs formed the derivation and validation datasets. For models that used the expert-selected predictors, imputation was conducted and five imputed datasets were created for derivation and validation datasets each. In terms of analyses, C-index, AUPRC, calibration, net benefit, and positive/ negative capture analyses were all conducted on the validation set. For the training of TRisk and DeepSurv models, to avoid overfitting, 5% of the derivation dataset was randomly selected for end-of-epoch testing (i.e., end-of-epoch test dataset). After training converged (i.e., the model testing loss converged and no longer reduced), the model was evaluated on the validation dataset. Full derivation dataset was used for fitting the conventional statistical models.

#### EHR pre-processing and modelling of TRisk

For TRisk modelling, we mapped all recorded diagnoses in Read code from general practices to the ICD-10 code to preserve a consistent coding scheme between the records in the hospital and the general practices. The mapping relied on a phenotyping dictionary provided by NHS ­­digital and SNOMED-CT. Additionally, the medications were recorded in the format of the British National Formulary (BNF) coding scheme, and we kept the codes in the section level (four digits). Furthermore, the procedure and lab test were recorded as OPCS codes and Read codes respectively, and we retained those codes in the original format.

To explain the transformation from BEHRT into TRisk, the BEHRT model is first described. The model, BEHRT, has three components: (1) An embedding layer that incorporates three layers of raw EHR data for modelling (see **Figure S1**), (2) the Transformer-based feature extractor that handles data and extracts features from the temporal EHR data that are useful for the prediction task and (3) the binary prediction layer, which employs a sigmoid activation function following a linear transformation of the patient representation latent state to yield risk estimates (i.e., probability of occurrence of outcome) for a given patient.^1^ Since BEHRT is a binary classification model, the objective function is the binary cross entropy loss function.

BEHRT is a Transformer-based sequential deep learning model that incorporates four layers of features for risk prediction. The features include encounter, age, segmentation, and positional encoding. Encounter comprises diagnoses, medications, and other available information in the EHR dataset that represents an individual’s medical trajectory prior to baseline. Age, segmentation, and positional encoding present the time-related information for each encounter to preserve the temporal order of each encounter. To be more precise, "age" refers to the specific age recorded for an encounter that took place. (calculated using event date and the date of birth), segmentation are two symbolic embeddings change alternatively between visits to distinguish latent representation of encounter in different visits. The positional encoding is pre-calculated embeddings that provide sequential information for the Transformer model. The summation of the embeddings from all four features is the final representation of a corresponding encounter. Afterwards, the Transformer model is used to extract the temporal interactions and latent features for the EHR sequence, and we used the first-time step of the final layer of Transformer as the patient representation for CVD risk prediction.

In gist, the transformation of BEHRT into the TRisk survival model involved substituting (3) the binary cross-entropy loss function with a survival modelling-driven loss function, while retaining the sophisticated (2) feature extraction capabilities of BEHRT (i.e., the Transformer layers).^1^ Lastly, we also augmented (1) the embedding structure with respect to original BEHRT architecture.

Starting with the embedding layers, first, the age embedding now explicitly includes age at baseline to more expressively capture time of prediction. In this way, for a particular patient, prediction at age, 61 years, will be different than, say at age, 65 years, and hence will be appropriately denoted differently. Also, in this way, baseline age is explicitly included as a variable for use in prediction of the outcome of interest.

Also, the embedding layers are not summed like in BEHRT.^1^ Rather, the three separate embedding layers are concatenated using tensor concatenation operations and multiplied by a weight vector that transforms the space: 3•E → E (i.e., E being the space of one embedding layer; 3E is three stacked layers). Finally, a non-linear, hyperbolic tangent functional transformation is applied to the product to enable more expressive latent representation of input raw EHR and ultimately, mitigate overfitting.

In terms of the outcome modelling module of the TRisk model, the sigmoid activation and binary cross entropy objective function is replaced with the Scalable Continuous-Time Survival Model through Ordinary Differential Equation Networks (SODEN) framework for survival modelling. Instead of maximum likelihood estimation (MLE) frameworks that utilise costly integral calculations for model training on censored data, the SODEN framework alternatively poses MLE as a differential-equation constrained optimisation objective.^2^ Furthermore, unlike previous proportional hazard frameworks (e.g., DeepSurv model^3^) that have not been shown to be theoretically robust for stochastic gradient descent (SGD) using mini-batching (i.e., random non-overlapping subsets of the derivation dataset), the SODEN framework alleviates many issues of established survival model frameworks that have trouble scaling to “big data”. Specifically, with ordinary differential equations modelling the time-to-event distribution, the framework for MLE on censored data becomes more flexible (i.e., absent of strong structural assumptions of the shape of the survival/hazard distribution), more appropriately scalable (i.e., can be trained using SGD).

Bottom of FormFinally, the TRisk objective or cost function utilises an Explicit Calibration (EC) method for optimising for D-calibration, an omnibus measure of calibration, in addition to empirical loss.^4^

The TRisk model similar to the BEHRT model utilises the prediction token (‘Predict token’) to serve as a dense patient representation vector for each patient. This vector functioning as a dense feature vector is then fed into the SODEN framework survival modelling network that conducts risk prediction for a particular outcome. With these modular components, the TRisk model is theoretically more robust for scalable training on large-scale EHR data than other DL survival modelling frameworks and ensures greater expressiveness and flexibility for improved discrimination and calibration.

#### Predictor selection for benchmark models

The raw variables extracted for locally fitted SCORE2 CPH modelling included sex, age, smoking status, diabetes, systolic blood pressure, total cholesterol, and high-density lipoprotein cholesterol. In addition to these variables, the final SCORE2 model included age interactions for all predictors except sex. All processing for the variables, including imputation, was conducted as in the validation study by the SCORE2 working group^5^. In brief, predictors were captured from both primary care and HES records. Missing values for systolic blood pressure, total cholesterol, and high-density lipoprotein cholesterol were imputed (more details on imputation below).

For the sex-specific QRISK3 modelling and DeepSurv modelling, the predictors included age, ethnicity, systolic blood pressure, standard deviation of systolic blood pressure, body mass index, total cholesterol to high-density lipoprotein cholesterol ratio, smoking status, family history of coronary heart disease in a first degree relative aged under 60 years, diabetes (type 1 and 2), rheumatoid arthritis, atrial fibrillation, chronic kidney disease (stage 3, 4, and 5), migraine, lupus erythematosus, severe mental illness, HIV or AIDS, erectile dysfunction, treated hypertension, history of prescribing of atypical antipsychotic drugs, and corticosteroid use. As our CPRD data did not provide Townsend score variables, we did not include it in the modelling. All processing for the variables was conducted as described in the validation study by Hippisley-Cox et al.; the appropriate fractional polynomial and interaction terms were modelled using source code from the ClinRisk website (<https://qrisk.org/src.php>). The following variables underwent imputation: smoking status, systolic blood pressure, standard deviation of systolic blood pressure, body mass index, total cholesterol, and high-density lipoprotein cholesterol (more details on imputation below).

Missing values for the QRISK3 and SCORE2 predictors were: smoking status (48% missing in general population cohort; 13% missing in the diabetes cohort), systolic blood pressure (39%; 3%), standard deviation of systolic blood pressure (64%; 9%), body mass index (59%; 10%), total cholesterol (73%; 6%), and high-density lipoprotein cholesterol (81%; 22%).

For the benchmark models, we extracted systolic blood pressure, body mass index, total cholesterol, and high-density lipoprotein cholesterol measurements using mean value within the range of maximum 2 years before the baseline and identified the prevalence of diagnoses using phenotyping provided by the CALIBER repository and medications using the British National Formulary codes.

#### Survival modelling

The locally fitted survival models (i.e., the SCORE2 models) were implemented within the Cox proportional hazard (CPH) framework. This means the survival models included two components: the non-parametric baseline survival function $\lambda_{0}\left( t \right)$ and the partial hazard $e^{h\left( x \right)}$. We denote the $h\left( x \right)$ as the log-risk function. Therefore, the hazard function can be formulated as the following:

$$\lambda\left( t | x \right)= \lambda_{0}e^{h(x)}$$

For the conventional CPH model such as SCORE2, the log-risk function is a linear regression model. For the DeepSurv model, the linear log-risk structure is replaced with a multi-layer perceptron model^3^ and a linear activation function for the output layer to be trained with the partial log-likelihood objective function.

#### Implementation details: general population cohort

Implementation of statistical models and imputation details

Imputation of missing variables in addition to the implementation and analyses of the QRISK3 model and locally fitted SCORE2 CPH models were carried out in R and Python programming languages. Imputation for QRISK3 was carried out as described by derivation study; particularly, age interaction terms, the Nelson-Aalen estimator of the baseline cumulative hazard, and the outcome variable (i.e., incident CVD) were included for the multiple imputation process.^6^

In total, for each model, five imputations were conducted on each derivation and validation datasets of general population cohort using the “mice: Multivariate Imputation by Chained Equations” package in R.

After doing so, the implementation of the QRISK3 model was carried out using codes developed by ClinRisk Ltd. The C codes were extracted from the QRISK3 online score calculator website (<https://qrisk.org/src.php>) and converted to python codes for running on each imputed validation dataset (5 imputed validation datasets). Then, the predictions on each imputed validation dataset were pooled and collected for downstream analysis using appropriate extensions of Rubin’s rules (i.e., using complementary log-log transformations).

For locally fitted SCORE2 CPH modelling, the models were fit on each of the five imputed derivation datasets with appropriate predictors (i.e., 5 SCORE2 models for general population cohort modelling). The models were pooled using Rubin’s rules and evaluated on the 5 imputed validation datasets; predictions were pooled and collected for downstream analysis using appropriate extensions of Rubin’s rules (i.e., using complementary log-log transformations).

Implementation of deep learning models

For imputation of variables for utilisation in DeepSurv, as it is a DL model, the same imputed datasets as were used for QRISK3 were used for DeepSurv. After training five models (as there are five imputed datasets), however, model pooling could not be implemented as conventional pooling strategies developed for the conventional statistical framework could not be transferred to the DL framework. Rather, we extracted predictions from each of the respective models for each imputed dataset and used appropriate extensions of Rubin’s rules for prediction pooling (i.e., using complementary log-log transformations).

In terms of fitting of DeepSurv models on large-scale EHR, while the partial log-likelihood objective function is an appropriate adaptation of the proportional hazard framework to the non-linear DL setting, the function requires access to the entire dataset to identify the order of the positive cases for loss calculation. Therefore, it may fail when the dataset is too large to fit in the memory or if the model is too computationally expensive to fit. We have trained the model with mini-batch training as is conventional for this type of model; this is a stochastic approximation technique for model training and the model can converge similarly as training on the entire dataset. In terms of the implementation, the method is identical to the stochastic gradient descent, and random sampling can be achieved by shuffling the dataset every epoch.

On the other hand, there is no such issue when training the SODEN-framework TRisk model. The TRisk model does not require baseline hazards computation as well. TRisk is trained in a conventional DL end-to-end fashion; testing and evaluation on validation dataset just requires estimating appropriate hazard curves at the individual level at the 10-year mark.

In terms of the model training, TRisk and DeepSurv were trained using the Adam optimiser. Additionally, while the TRisk were trained with a Cosine annealing learning rate scheduler, the DeepSurv implementations were trained with a fixed learning rate.^7^ In this work, TRisk and DeepSurv were trained using the Pytorch coding framework for DL model development.

In terms of the two DL models’ hyperparameter selection and optimisation, we derived a train and test dataset from the derivation dataset of the general population cohort (i.e., the dataset with 2.2 million patients) in order to find the optimal hyperparameters that maximise performance in terms of C-index. For each DL model, we randomly selected 80% of patients in the derivation dataset of the general population cohort, trained the model for 100 epochs, and enabled early stopping if the testing loss on the test set (i.e., the remaining 20% of the patients) did not decrease for 10 epochs. A random search over 30 sets of hyperparameters for DeepSurv model was conducted. We adopted hyperparameters for TRisk from previously reported research^8^. The hyperparameter sets and optimal values for TRisk and DeepSurv models are shown in **Tables S1 and S2** respectively.

#### Modelling in diabetes cohort

For benchmark models, as imputation was already conducted for the derivation and validation datasets for the general population cohort, the SCORE2 and DeepSurv benchmark models were derived and validated on patients with diabetes with variables and processing as described previously. QRISK3 was applied on the validation dataset of the diabetes cohort. The same procedures for model and prediction pooling as described for the general population cohort was conducted for the diabetes cohort. The same hyperparameters used for DeepSurv used in the general population cohort was utilised for derivation of DeepSurv model in the diabetes cohort.

Transfer learning framework for TRisk

Transformer-based DL models such as TRisk require a large dataset for model training, which can be challenging to achieve when only a relatively small subset of the population is available, such as the subgroup with diabetes in our study. To overcome this challenge, we conducted a form of transfer learning and finetuned the TRisk model trained originally on the general population, on the subset of patients with diabetes.^7^ More specifically, after transferring the model trained in the general population cohort, we only finetuned the pooling and SODEN prediction layers of TRisk with all weights of the previous Transformer layers frozen (i.e., fixed and not trainable).

### Supplementary Results

#### Impact analysis

The impact analysis of models at different risk thresholds and standardised to a population size of 1000 is shown in Table 2. In the general population cohort, our impact analysis demonstrated that applying QRISK3 at the commonly recommended 10% risk threshold would classify 203,154 patients (27% of population) as eligible for treatment with 26,314 patients with an event (13% true positive) over the 10-year follow-up. The remaining 543,922 patients (73% of population) were ranked as low-risk of whom 536,988 (~99%) were patients free of event (i.e., true negatives). Raising the threshold to 15% would restrict the cohort of eligible patients for preventative therapy to 19% of the population, but simultaneously increase false negatives from 6,934 to 11,290 patients.

In comparison, TRisk was not only better at stratifying risk as compared to QRISK3 at the 10% risk threshold but also, the model could outperform benchmarks at thresholds up to and including 15% without detrimental consequences. At the 15% illustrative threshold for TRisk, 133,405 patients (18% of cohort) would be eligible for treatment. Among those, 27,499 (21%) were those who indeed had CVD outcome (i.e., true positive patients). The remaining 613,671 patients (82% of population) were ranked as low-risk, with 607,922 (>99%) of those being true negatives and 1,185 fewer false negatives than QRISK3 at 10% threshold. At both the 10% and 15% illustrative thresholds, TRisk improves upon the recommended strategy: in the former, by delivering superior true positive capture and in the latter, by finer selection of those considered high-risk.

For the diabetes cohort, the TRisk approach similarly demonstrated better positive and negative capture as compared to conventional approaches. At the threshold of 10%, TRisk captured a more refined population of 10,999 (76% of population) patients with 1,667 (15%) patients with correctly identified cases of CVD (i.e., true positive patients). As compared to the conventional indiscriminate approach of recommending all patients with diabetes for treatment, TRisk would recommend 24% fewer patients with minimal false negative capture of 28 patients (0.2% of population).

### Supplementary Tables

**Table S1.** Model parameters for TRisk

|  | General population cohort |
| --- | --- |
| Number of layers | 6 |
| Maximum sequence length | 512 |
| Hidden size | 256 |
| Hidden dropout rate | 0.2 |
| Attention dropout rate | 0.2 |
| Number of attention heads | 4 |
| Intermediate size | 512 |
| Pooling layer size | 256 |
| Hidden activation function | gelu |
| Learning rate | 0.00005 |
| Weight decay | 0.02 |

**Table S2**. Model parameters for DeepSurv

| Number of layers | Neurons | Batch norm | Dropout rate | Learning rate |
| --- | --- | --- | --- | --- |
| 3 | 14, 14, 14 | True | 0.2 | 0.0001 |

**Table S3**: Area under the precision-recall curve (AUPRC) for the general population cohort

| Model | AUPRC |
| --- | --- |
| SCORE2* | 0.177 |
| QRISK3 | 0.184 |
| DeepSurv | 0.191 |
| TRisk | 0.346 |

*Locally fitted Cox model

**Table S4**: Model discrimination analysis in terms of C-index of subgroups by different age ranges for the general population cohort

| Model | Age group | C-index (95% CI) |
| --- | --- | --- |
| SCORE2* | 40-69 | 0.743 (0.735, 0.750) |
|  | 40-84 | 0.778 (0.773, 0.783) |
| QRISK3 | 40-69 | 0.730 (0.722, 0.738) |
|  | 40-84 | 0.768 (0.763, 0.773) |
| DeepSurv | 40-69 | 0.764 (0.765, 0.771) |
|  | 40-84 | 0.790 (0.785, 0.796) |
| TRisk | 40-69 | 0.884 (0.878, 0.889) |
|  | 40-84 | 0.874 (0.870, 0.879) |

95% CI: 95% confidence interval; *locally fitted Cox model

**Table S5**: Sex-stratified discrimination performance for the general population cohort

| Model | Sex | C-index (95% CI) |
| --- | --- | --- |
| SCORE2* | Male | 0.815 (0.809, 0.822) |
|  | Female | 0.851 (0.844, 0.858) |
| QRISK3 | Male | 0.809 (0.803, 0.816) |
|  | Female | 0.848 (0.841, 0.855) |
| DeepSurv | Male | 0.827 (0.820, 0.833) |
|  | Female | 0.859 (0.852, 0.865) |
| TRisk | Male | 0.896 (0.891, 0.901) |
|  | Female | 0.922 (0.917, 0.927) |

95% CI: 95% confidence interval; *locally fitted Cox model

**Table S6**: Discrimination performance stratified by levels of index of multiple deprivation (IMD) for the general population cohort

| Model | IMD | C-index (95% CI) |
| --- | --- | --- |
| SCORE2* | 1 | 0.845 (0.835, 0.855) |
|  | 2 | 0.840 (0.830, 0.851) |
|  | 3 | 0.837 (0.827, 0.847) |
|  | 4 | 0.833 (0.822, 0.843) |
|  | 5 | 0.830 (0.818, 0.842) |
| QRISK3 | 1 | 0.837 (0.827, 0.848) |
|  | 2 | 0.834 (0.823, 0.844) |
|  | 3 | 0.829 (0.819, 0.839) |
|  | 4 | 0.826 (0.815, 0.836) |
|  | 5 | 0.823 (0.820, 0.835) |
| DeepSurv | 1 | 0.851 (0.841, 0.862) |
|  | 2 | 0.849 (0.839, 0.859) |
|  | 3 | 0.843 (0.833, 0.853) |
|  | 4 | 0.839 (0.829, 0.849) |
|  | 5 | 0.837 (0.826, 0.849) |
| TRisk | 1 | 0.914 (0.906, 0.922) |
|  | 2 | 0.911 (0.903, 0.919) |
|  | 3 | 0.909 (0.901, 0.917) |
|  | 4 | 0.906 (0.898, 0.915) |
|  | 5 | 0.905 (0.895, 0.914) |

95% CI: 95% confidence interval; *locally fitted Cox model

**Table S7.** Population characteristics for derivation and validation datasets of diabetes cohort

|  | | Derivation  (n=44,668) | Validation  (n=14,518) |
| --- | --- | --- | --- |
| CVD cases (%) | | 5,702 (12.77) | 1,695 (11.68) |
| Women (%) | | 20,232 (45.29) | 6,556 (45.16) |
| Mean age in years (SD) | | 60 (15) | 59 (15) |
| Ethnicity | | | |
| Unknown (%) | | 27,190 (60.87) | 8,909 (61.37) |
| White (%) | | 15,646 (35.03) | 5,008 (34.5) |
| Other Asian (%) | | 204 (0.46) | 60 (0.41) |
| Pakistani (%) | | 293 (0.66) | 65 (0.45) |
| Indian (%) | | 397 (0.89) | 118 (0.81) |
| Other (%) | | 228 (0.51) | 87 (0.60) |
| Caribbean (%) | | 209 (0.47) | 119 (0.82) |
| Mixed (%) | | 81 (0.18) | 22 (0.15) |
| Bangladeshi (%) | | 105 (0.24) | 26 (0.18) |
| Chinese (%) | | 56 (0.13) | 15 (0.10) |
| Other Black (%) | | 90 (0.20) | 29 (0.20) |
| Black African (%) | | 169 (0.38) | 60 (0.41) |
| Index of multiple deprivation (IMD) | | | |
| IMD 1 (%) | | 8,100 (18.13) | 2,612 (17.99) |
| IMD 2 (%) | | 9,307 (20.84) | 2,860 (19.7) |
| IMD 3 (%) | | 9,305 (20.83) | 3,125 (21.53) |
| IMD 4 (%) | | 9,112 (20.40) | 3,163 (21.79) |
| IMD 5 (%) | | 8,844 (19.80) | 2,758 (19.00) |
| Region | | | |
| North East (%) | | 539 (1.21) | 586 (4.04) |
| North West (%) | | 7,403 (16.57) | 1,206 (8.31) |
| Yorkshire and the Humber (%) | | 1,395 (3.12) | 694 (4.78) |
| East Midlands (%) | | 1,290 (2.89) | 776 (5.35) |
| West Midlands (%) | | 4,823 (10.80) | 1,781 (12.27) |
| East of England (%) | | 5,261 (11.78) | 1,328 (9.15) |
| South West (%) | | 5,085 (11.38) | 2,684 (18.49) |
| South Central (%) | | 5,897 (13.20) | 1,280 (8.82) |
| London (%) | | 7,327 (16.40) | 2,304 (15.87) |
| South East Coast (%) | | 5,648 (12.64) | 1,879 (12.94) |
| Mean systolic blood pressure^†^, mmHg (SD) | | 136.65 (14.23) | 136.45 (14.16) |
| Mean body mass index^†^, kg/m^2^ (SD) | | 30.65 (5.89) | 30.72 (5.91) |
| Mean high density lipoprotein^†^, mmol/L (SD) | | 1.23 (0.40) | 1.24 (0.55) |
| Mean total cholesterol^†^, mmol/L (SD)^†^ | | 4.7 (1.83) | 4.74 (1.49) |
| Smoking status^†^ (number of cigarettes per day) | | | |
| Non-smoker (%) | | 22,627 (50.66) | 7,371 (50.77) |
| Ex-smoker (%) | | 14,880 (33.31) | 4,752 (32.73) |
| Light (<10 cigarettes/day) (%) | | 1,936 (4.33) | 679 (4.68) |
| Moderate (10-20 cigarettes/day) (%) | | 2653 (5.94) | 861 (5.93) |
| Heavy (>20 cigarettes/day) (%) | | 2,572 (5.76) | 855 (5.89) |
| Comorbidities at baseline | | | |
| Diabetes (%) | | 44,668 (100.00) | 14,518 (100.00) |
| Rheumatoid arthritis (%) | | 375 (0.84) | 133 (0.92) |
| Atrial fibrillation (%) | | 1,795 (4.02) | 552 (3.80) |
| CKD (%) | | 1,774 (3.97) | 597 (4.11) |
| Migraine (%) | | 1,057 (2.37) | 349 (2.40) |
| Lupus erythematosus (%) | | 43 (0.10) | 14 (0.10) |
| Mental illness (%) | | 678 (1.52) | 239 (1.65) |
| HIV/AIDS (%) | | 138 (0.31) | 50 (0.34) |
| Erectile dysfunction (%) | | 4,050 (9.07) | 1,299 (8.95) |
| Medication use at baseline | | | |
| Antihypertensives (%) | | 3,640 (8.15) | 1,096 (7.55) |
| Antipsychotics (%) | | 125 (0.28) | 32 (0.22) |
| Corticosteroids (%) | | 575 (1.29) | 162 (1.12) |

SD: standard deviation; %: percentage; CVD: cardiovascular disease; IMD: Index of multiple deprivation; CKD: chronic kidney disease;

^†^: missing observations: smoking status (13% missing in the diabetes cohort, systolic blood pressure (3%), standard deviation of systolic blood pressure (9%), body mass index (10%), total cholesterol (6%), and high-density lipoprotein cholesterol (22%).

**Table S8**: Area under the precision-recall curve (AUPRC) for the diabetes cohort

|  | AUPRC |
| --- | --- |
| SCORE2* | 0.203 |
| QRISK3 | 0.223 |
| DeepSurv | 0.214 |
| TRisk | 0.323 |

*Locally fitted Cox model

**Table S9**: Model discrimination analysis in terms of C-index of subgroups by different age ranges for the diabetes cohort

| Model | Age group | C-index (95% CI) |
| --- | --- | --- |
| SCORE2* | 40-69 | 0.627 (0.588, 0.665) |
|  | 40-84 | 0.665 (0.639, 0.692) |
| QRISK3 | 40-69 | 0.639 (0.600, 0.678) |
|  | 40-84 | 0.672 (0.646, 0.698) |
| DeepSurv | 40-69 | 0.660 (0.622, 0.698) |
|  | 40-84 | 0.683 (0.657, 0.709) |
| TRisk | 40-69 | 0.780 (0.746, 0.814) |
|  | 40-84 | 0.771 (0.748, 0.795) |

95% CI: 95% confidence interval; *locally fitted Cox model

**Table S10**: Sex-stratified discrimination performance for the diabetes cohort

| Model | Sex | C-index (95% CI) |
| --- | --- | --- |
| SCORE2* | Male | 0.669 (0.635, 0.703) |
|  | Female | 0.711 (0.672, 0.751) |
| QRISK3 | Male | 0.669 (0.635, 0.703) |
|  | Female | 0.716 (0.677, 0.755) |
| DeepSurv | Male | 0.689 (0.655, 0.722) |
|  | Female | 0.722 (0.683, 0.761) |
| TRisk | Male | 0.776 (0.746, 0.807) |
|  | Female | 0.803 (0.768, 0.837) |

95% CI: 95% confidence interval; *locally fitted Cox model

**Table S11**: Discrimination performance stratified by levels of index of multiple deprivation (IMD) for the diabetes cohort

| Model | IMD | C-index (95% CI) |
| --- | --- | --- |
| SCORE2* | 1 | 0.681 (0.620, 0.743) |
|  | 2 | 0.717 (0.660, 0.774) |
|  | 3 | 0.695 (0.641, 0.749) |
|  | 4 | 0.661 (0.604, 0.718) |
|  | 5 | 0.696 (0.638, 0.754) |
| QRISK3 | 1 | 0.687 (0.626, 0.749) |
|  | 2 | 0.725 (0.669, 0.782) |
|  | 3 | 0.710 (0.656, 0.763) |
|  | 4 | 0.660 (0.603, 0.716) |
|  | 5 | 0.689 (0.63, 0.747) |
| DeepSurv | 1 | 0.680 (0.619, 0.742) |
|  | 2 | 0.747 (0.692, 0.802) |
|  | 3 | 0.718 (0.665, 0.771) |
|  | 4 | 0.674 (0.618, 0.730) |
|  | 5 | 0.694 (0.636, 0.752) |
| TRisk | 1 | 0.773 (0.717, 0.829) |
|  | 2 | 0.796 (0.744, 0.847) |
|  | 3 | 0.808 (0.762, 0.855) |
|  | 4 | 0.766 (0.714, 0.817) |
|  | 5 | 0.794 (0.742, 0.846) |

95% CI: 95% confidence interval; *locally fitted Cox model

### Supplementary Figures

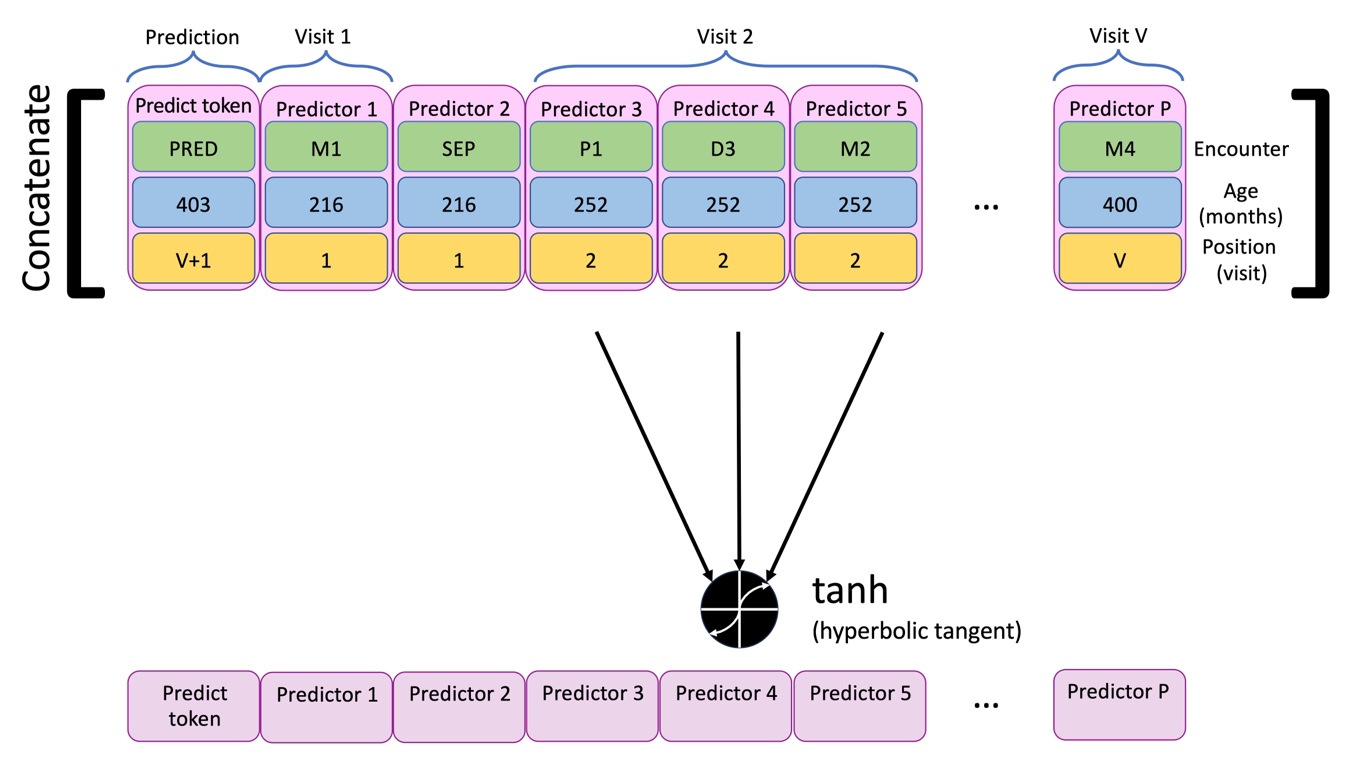

**Figure S1. TRisk input space for a hypothetical patient.** For a hypothetical patient, the figure presents a sequence of medical history records represented by a group of three variables. Each record is comprised of an encounter code (e.g., “M1”), the age at which the encounter was recorded in years (e.g., “216” months), and the visit number (e.g., visit “1”). These three components of the raw electronic health record data are represented by embeddings inputted into the model. The first character of the encounter code, D, M, and P represent diagnosis, medication, and procedure records respectively represented by a code in this figure. The number following the letter illustrates a hypothetical code of that modality type (e.g., ‘D3’ represents code “I51.2” in ICD-10 encoding). “SEP” represents a separation character given to the model to split up the data between visits. Lastly, the “PRED” token (i.e., “predict token”) is a special token that is used for prediction of the outcome (e.g., cardiovascular disease prediction). This special token has the age at baseline (e.g., age of “403” months at the time of index date/baseline for this hypothetical patient). As this is the visit that will happen following the final visit prior to baseline (i.e., visit number “V”), the visit for the predict token is appropriately enumerated as visit number “V+1” signifying prediction at the baseline “visit”. Only data up to baseline is included in the model. This predict token is inputted as displayed and following further transformation by Transformer architecture layers, the corresponding output state token will be used as a condensed latent patient representation layer for input into the ordinary differential equation-based survival prediction network layers.

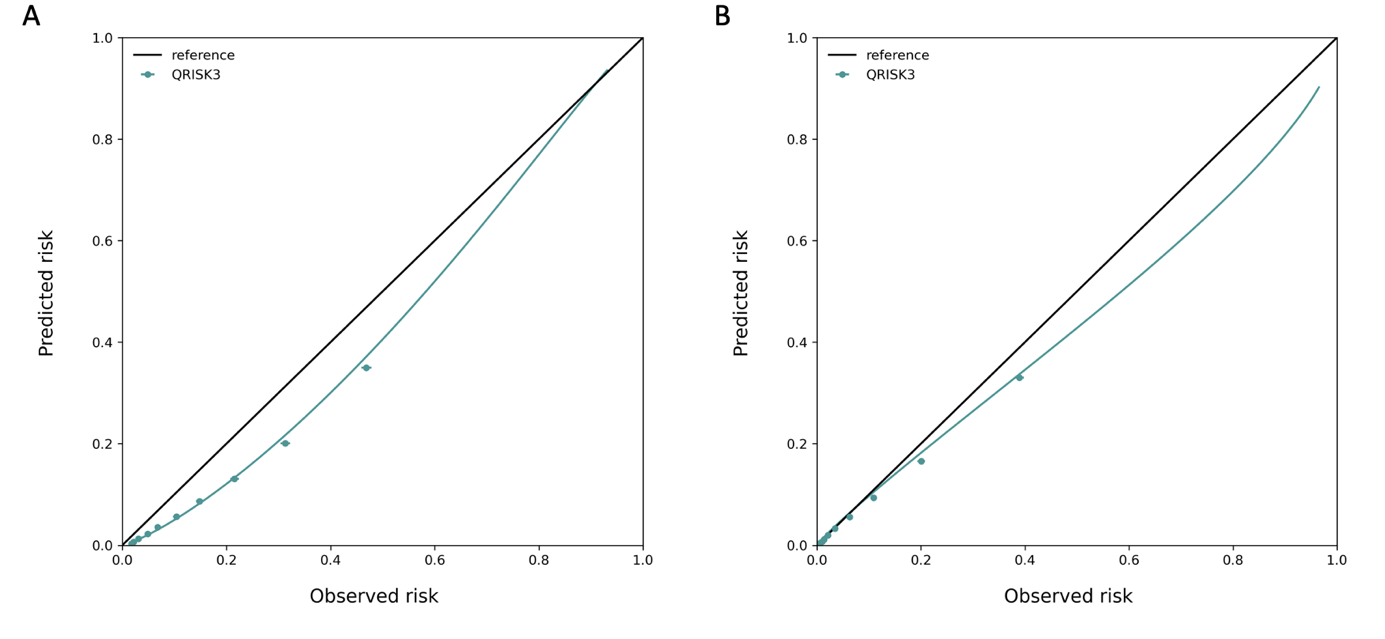

**Figure S2. QRISK3 sex-specific calibration plots in general population cohort.** Smoothed calibration lines and tenth of predicted risk decile calibration plots are shown for male (A) and female (B) sex-specific QRISK3 equations in general population cohort.

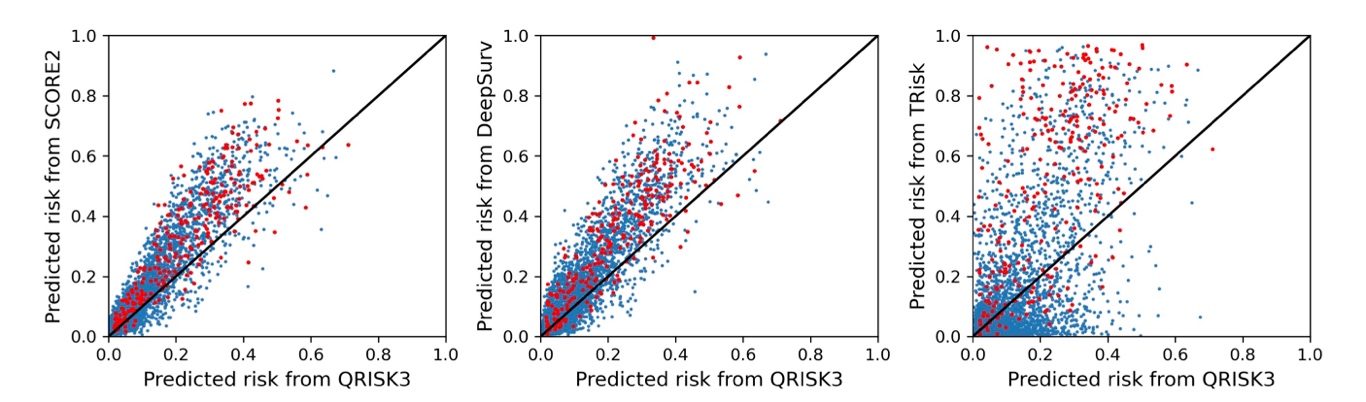

**Figure S3. Model consistency on individual level predictions in general population cohort.** QRISK3 is used as the benchmark model to compare with the SCORE2, DeepSurv, and TRisk in general population cohort. For the convenience of visualisation, only 500 patients were randomly selected for comparison. The red and blue dots represent patients with and without an event (or censored) in follow-up respectively.

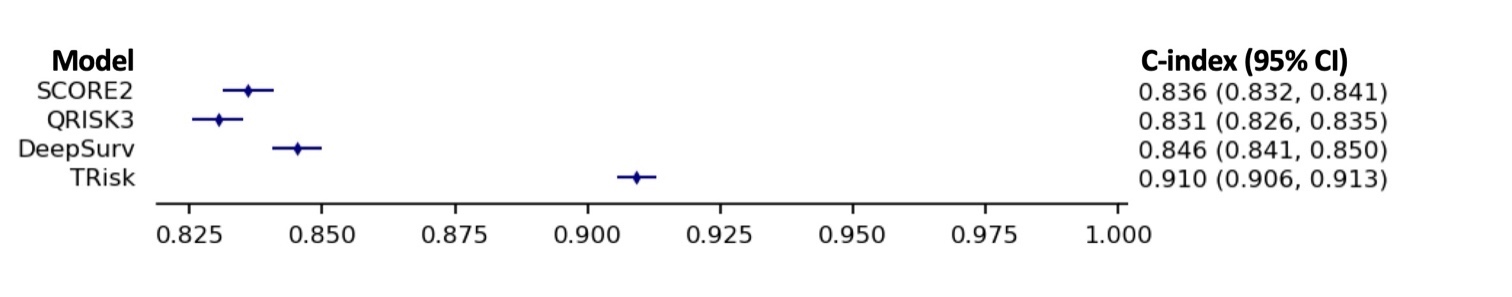

**Figure S4. Discriminative performance of models in general population cohort**. Discrimination is provided in this forest plot as assessed by C-index with 95% confidence intervals (CI) in general population cohort.

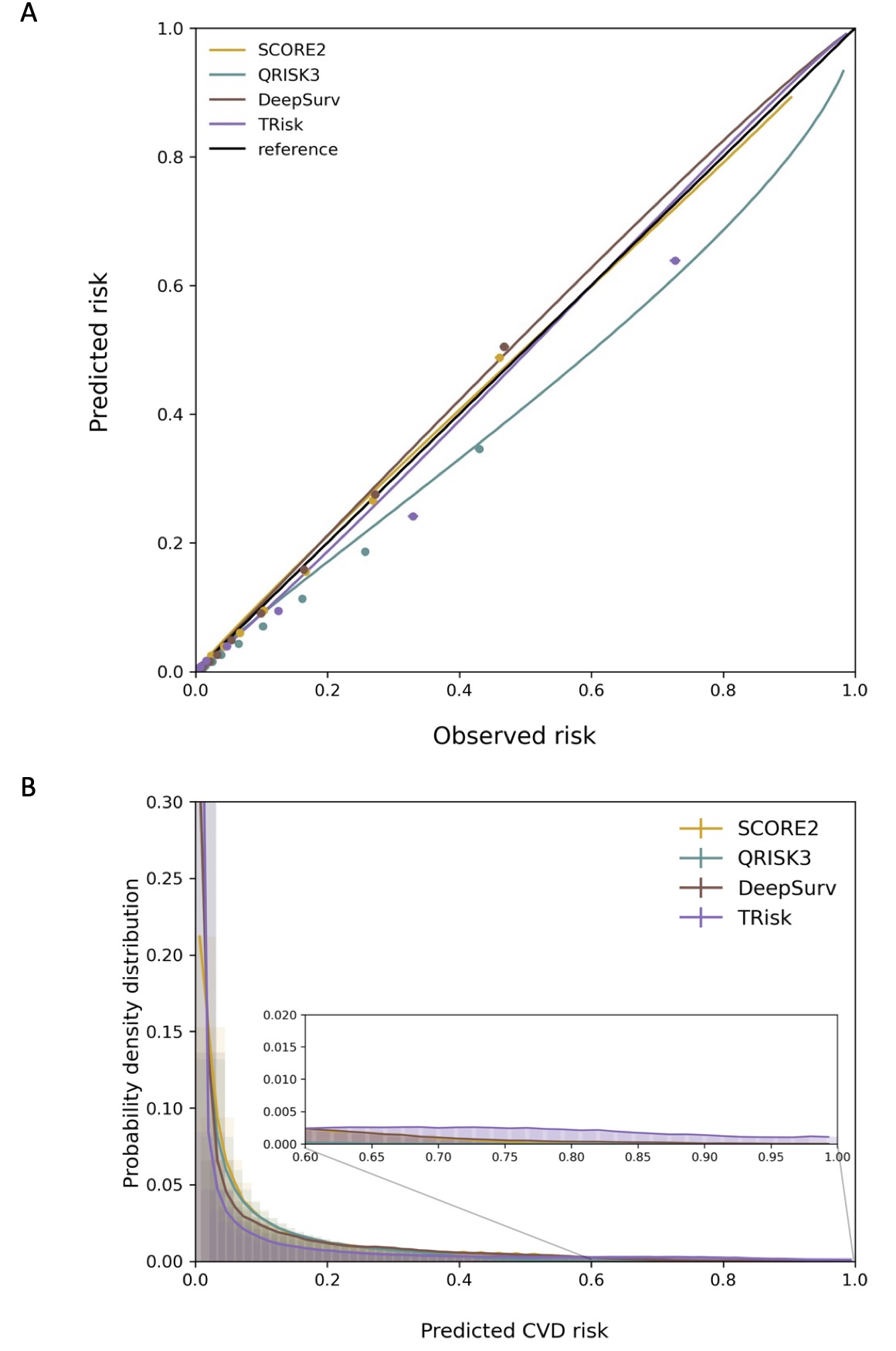

**Figure S5**. **Calibration plot and distribution of predicted risk of all models in general population cohort.** Smoothed calibration lines and tenth of predicted risk decile calibration plots (A) and distribution of predicted risk (B) are presented for all models implemented on general population cohort.

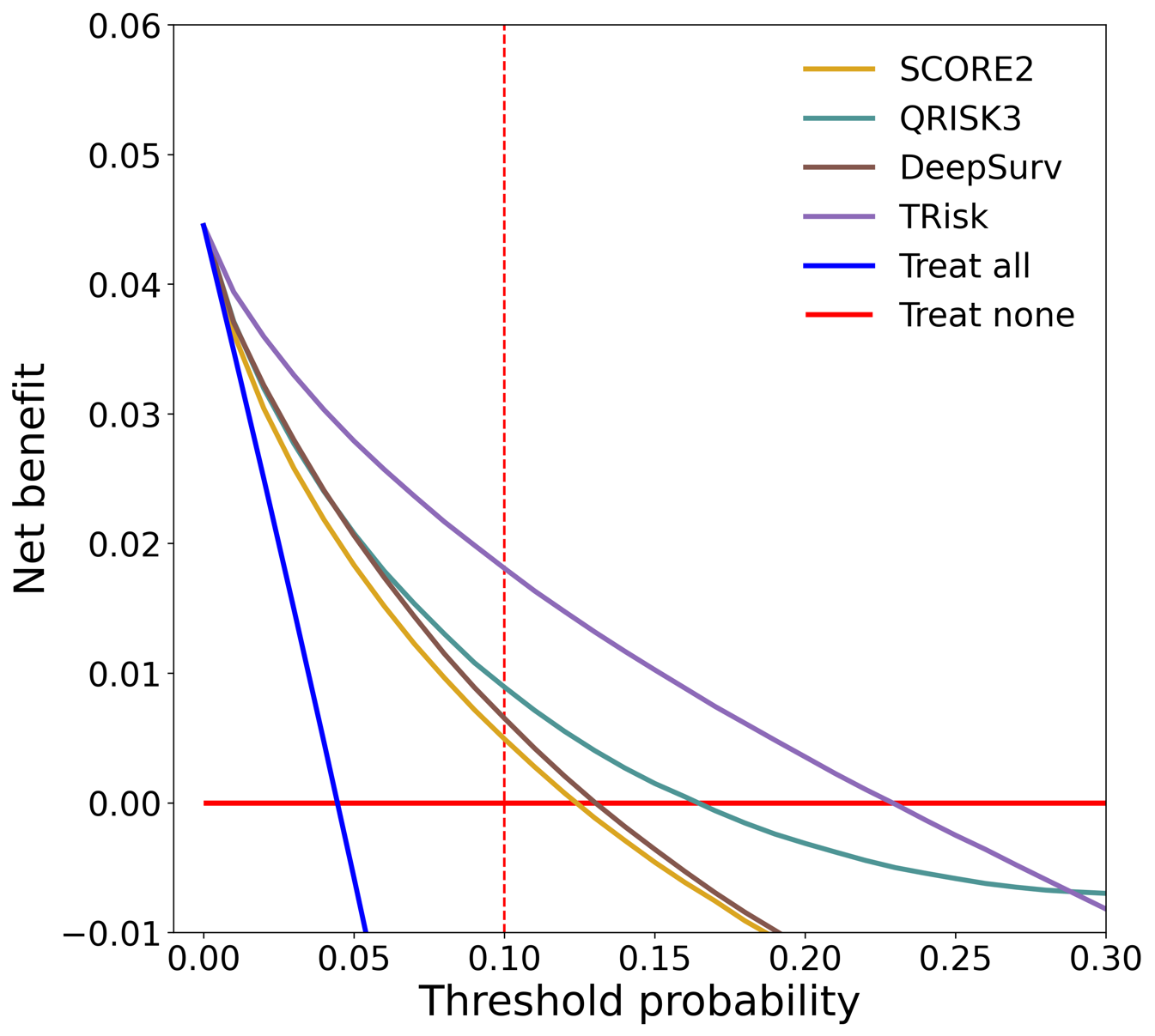

**Figure S6. Decision curve analysis for all models in general population cohort.** Decision curve analysis (including censored observations) is presented visually in this figure. Threshold probability is shown on the x-axis and the net benefit, a function of threshold probability, is shown on the y-axis and is the difference between the proportion of true positives and false positives weighted by odds of the respective decision threshold.

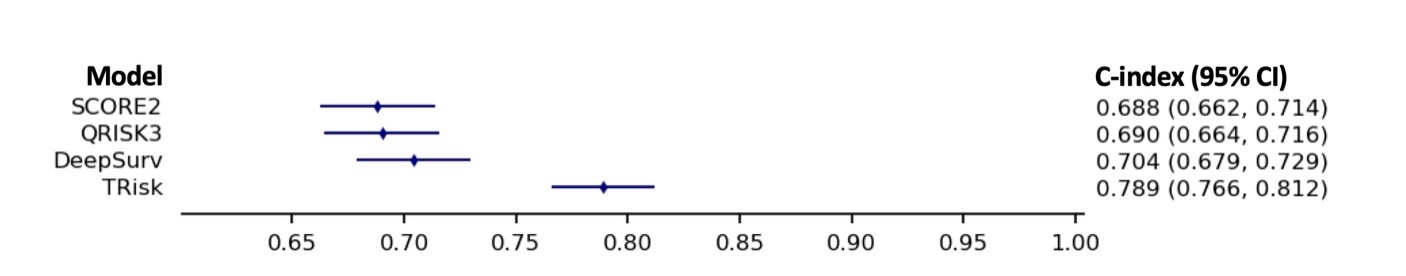

**Figure S7. Discriminative performance of models in diabetes cohort**. Discrimination is provided in this forest plot as assessed by C-index with 95% confidence intervals (CI) in diabetes cohort.

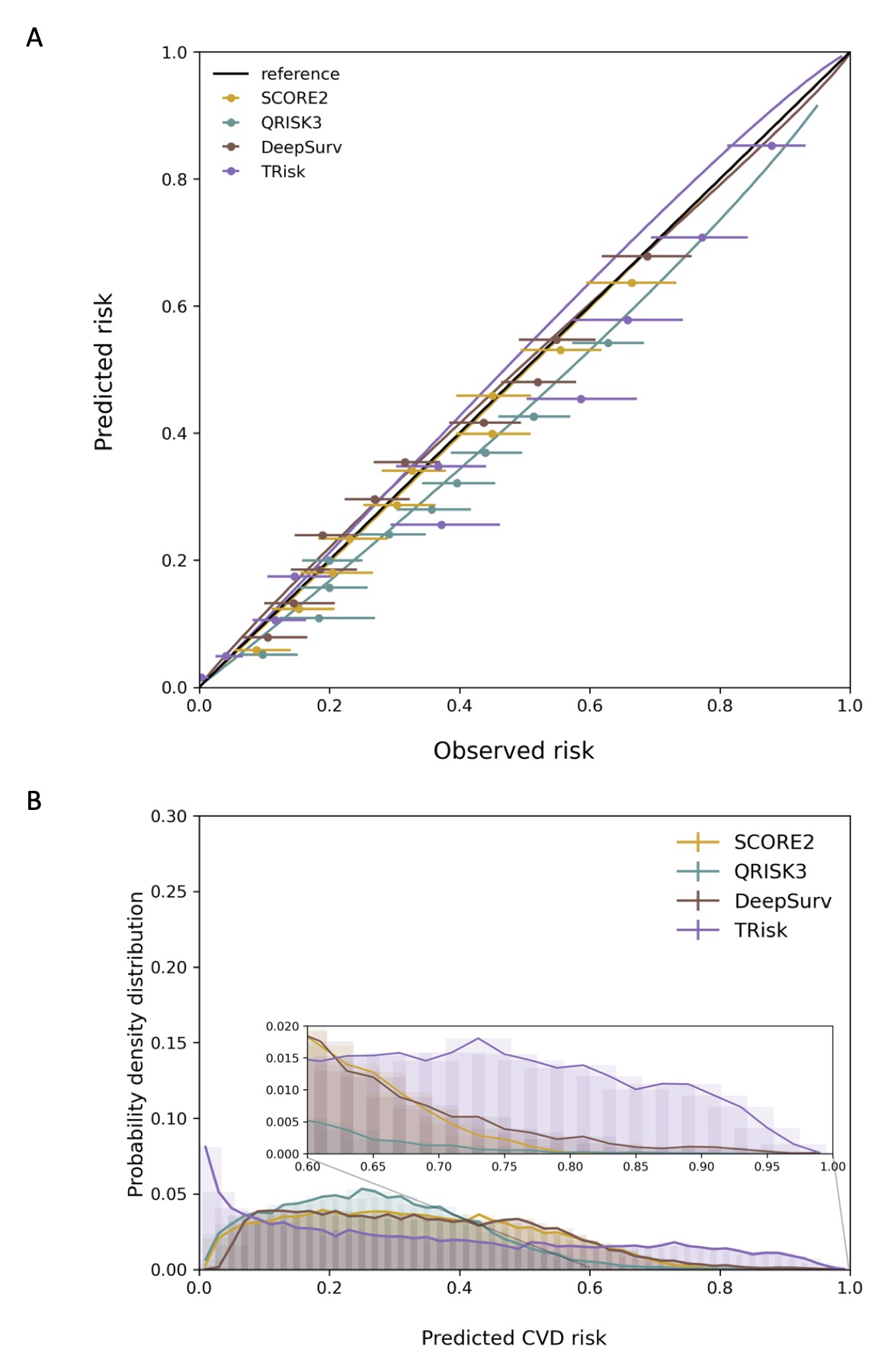

**Figure S8**. **Calibration plot and distribution of predicted risk of models in diabetes cohort.** Smoothed calibration lines and tenth of predicted risk decile calibration plots (A) and distribution of predicted risk (B) are presented for all models implemented on diabetes cohort.

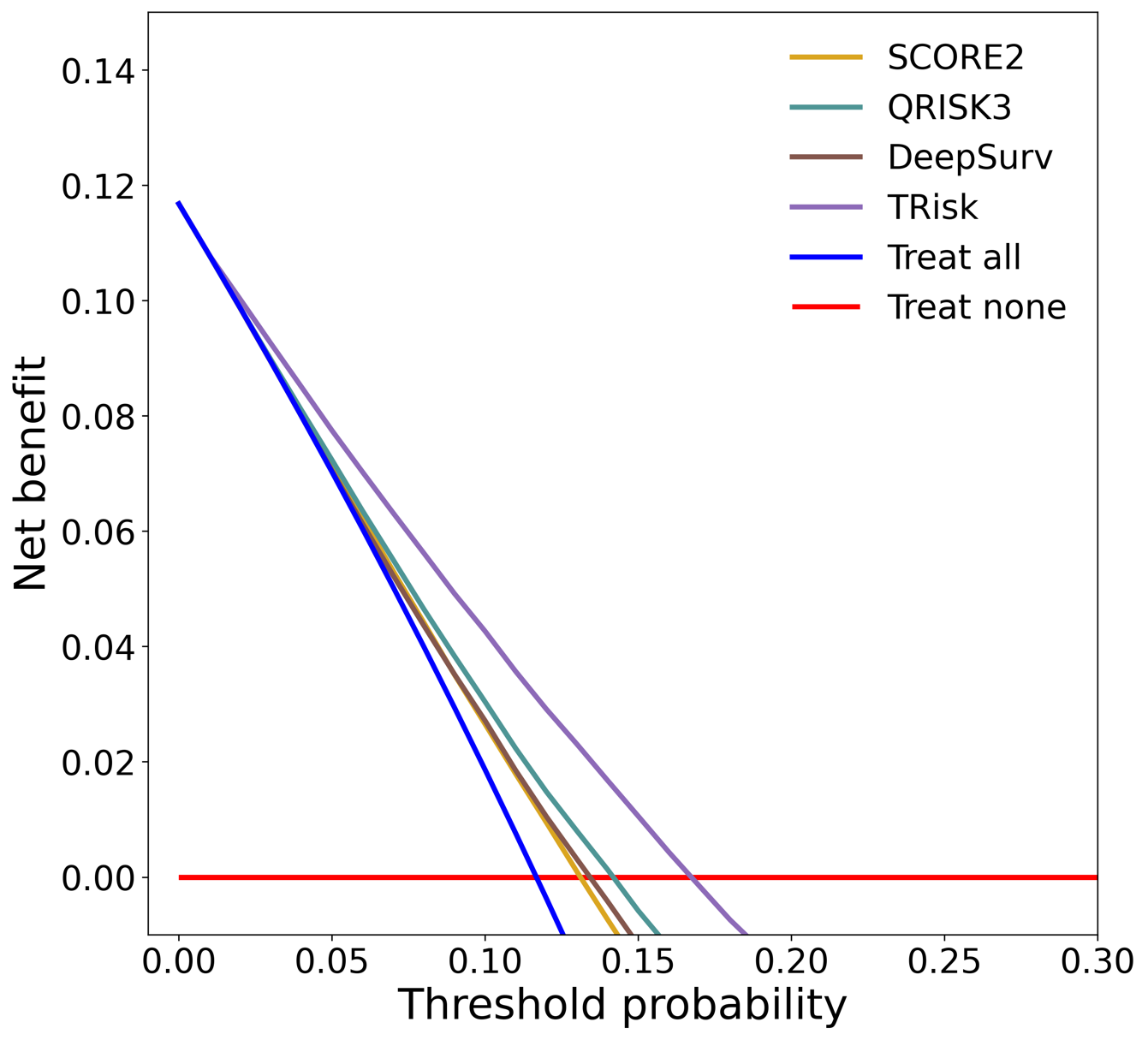

**Figure S9. Decision curve analysis for all models in diabetes cohort.** Decision curve analysis (including censored observations) is presented visually in this figure. Threshold probability is shown on the x-axis and the net benefit, a function of threshold probability, is shown on the y-axis and is the difference between the proportion of true positives and false positives weighted by odds of the respective decision threshold.

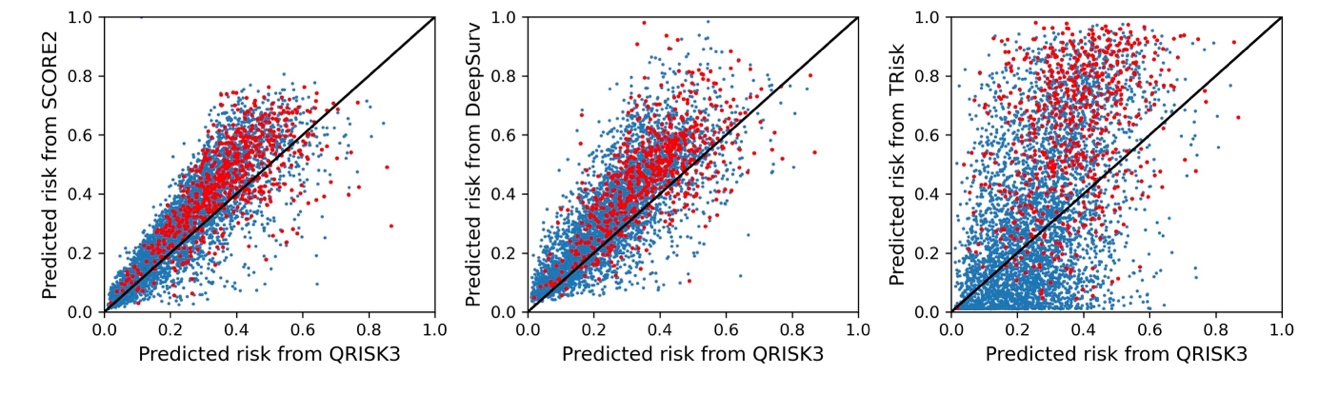

**Figure S10. Model consistency on individual level predictions in diabetes cohort.** QRISK3 is used as the benchmark model to compare with the SCORE2, DeepSurv, and TRisk on diabetes cohort. For the convenience of visualisation, only 500 patients were randomly selected for comparison. The red and blue dots represent patients with and without an event (or censored) in follow-up respectively.

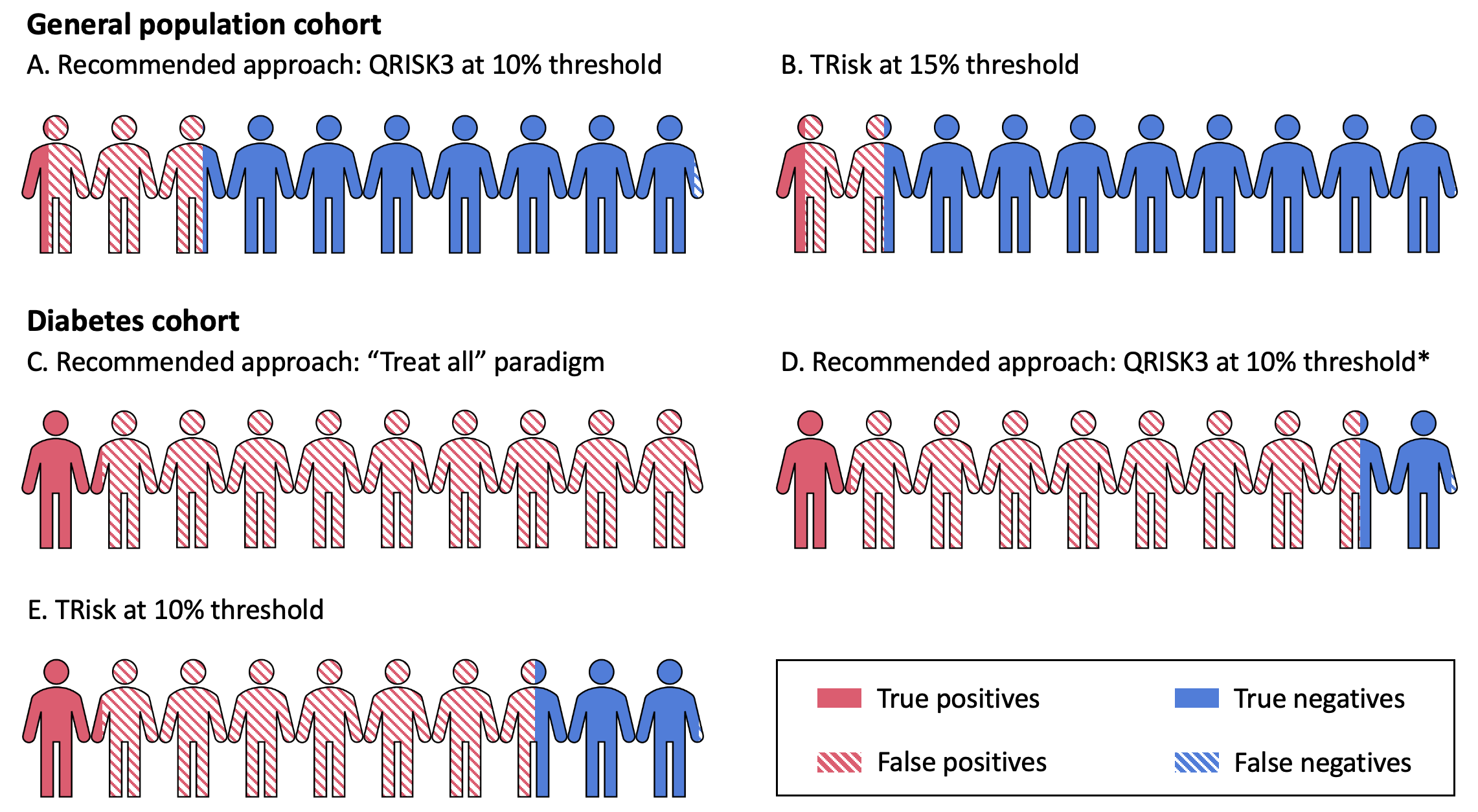

**Figure S11. Impact analysis summary.** For presentational purposes, hypothetical general population and diabetes cohorts of 10 patients are shown; the fraction of those coloured red and blue are true positives and negatives respectively with the shaded sections as false predictions. *Recently, UK guidelines have recommended a risk assessment strategy (i.e., QRISK3 at 10%) for preventative therapy allocation in patients with type 2 diabetes.

### References

1.	Li Y, Rao S, Solares JRA, Hassaine A, Ramakrishnan R, Canoy D, Zhu Y, Rahimi K, Salimi-Khorshidi G. BEHRT: Transformer for Electronic Health Records. *Sci Rep* 2020; 10: 7155.

2.	Tang W, Ma J, Mei Q, Zhu J. SODEN: A Scalable Continuous-Time Survival Model through Ordinary Differential Equation Networks, http://arxiv.org/abs/2008.08637 (2020).

3.	Katzman JL, Shaham U, Cloninger A, Bates J, Jiang T, Kluger Y. DeepSurv: Personalized treatment recommender system using a Cox proportional hazards deep neural network. *BMC Med Res Methodol*; 18.

4.	Goldstein M, Han X, Puli A, Perotte AJ, Ranganath R. X-CAL: Explicit calibration for survival analysis. In: *Advances in Neural Information Processing Systems*. 2020.

5.	SCORE2 risk prediction algorithms: New models to estimate 10-year risk of cardiovascular disease in Europe. *Eur Heart J*; 42. Epub ahead of print 2021. DOI: 10.1093/eurheartj/ehab309.

6.	Hippisley-Cox J, Coupland C, Brindle P. Development and validation of QRISK3 risk prediction algorithms to estimate future risk of cardiovascular disease: Prospective cohort study. *BMJ*; 357.

7.	Zhuang F, Qi Z, Duan K, Xi D, Zhu Y, Zhu H, Xiong H, He Q. A Comprehensive Survey on Transfer Learning, http://arxiv.org/abs/1911.02685 (2019).

8.	Li Y, Salimi-Khorshidi G, Rao S, Canoy D, Hassaine A, Lukasiewicz T, Rahimi K, Mamouei M. Validation of risk prediction models applied to longitudinal electronic health record data for the prediction of major cardiovascular events in the presence of data shifts. *European Heart Journal - Digital Health* 2022; ztac061.
